# Ribosomal DNA copy number variation shapes human physiology and disease risk

**DOI:** 10.64898/2026.01.09.26343685

**Authors:** Anil Raj, Jordan S. Brown, Nathaniel H. Thayer, Manuel Hotz, Irene Lam, Nicole Fong, Elena P. Sorokin, Marjola Thanaj, Daphna Rothschild, Jonathan K. Pritchard, Maria Barna, David G. Hendrickson

**Affiliations:** Calico Life Sciences LLC; Research Center for Optimal Health, School of Life Sciences, University of Westminster; Department of Genetics, Stanford University; Department of Biology, Stanford University

## Abstract

Variation in ribosomal DNA (rDNA) copy number influences diverse physiological traits in model organisms, yet its consequences for human health remain poorly characterized. Here, we provide the largest analysis of 45S rDNA copy number to date and the first population-scale characterization of 5S using whole-genome sequencing from 490,383 UK Biobank participants. Despite encoding components of the same molecular machine, these arrays vary independently and associate with divergent phenotypes. Higher 45S copy number associates with common metabolic diseases, increased adiposity, and hematological signatures reminiscent of ribosomopathies. Molecular characterization reveals a coherent through line: elevated secretory cell-derived proteins in plasma, altered proteostasis and translation gene programs across tissues, and increased glucose-stimulated insulin secretion in primary human pancreatic islets. In contrast, 5S copy number shows no disease associations but instead correlates with proportional organismal growth: increased lean mass and organ volumes. Here too, molecular signatures aligns with population-level findings, as tissue transcriptomics reveals changes to myogenic gene expression programs and altered fat metabolism. This multi-scale convergence establishes that the two human rDNA arrays function as independent genetic factors with divergent consequences for cellular physiology and human health.

## 2 Introduction

All eukaryotes maintain hundreds of ribosomal DNA (rDNA) copies in tandem arrays to meet cellular demands for ribosomal RNA, the essential components of the protein synthesis machinery [1–4]. In humans, rDNA exists in two tandem arrays: hundreds of 45S rDNA copies distributed across five acrocentric chromosomes, and approximately 100-200 copies of 5S rDNA in a single array on chromosome 1 [5,6]. The 45S array encodes the 18S, 5.8S, and 28S rRNAs through coordinated RNA Polymerase I transcription in the nucleolus, while 5S rRNA is transcribed separately by RNA Polymerase III in the nucleoplasm and participates in extra-ribosomal regulatory functions including ribosomal stress sensing via the RPL5-RPL11-p53 pathway [7–9]. Both arrays exhibit the repetitive architecture characteristic of multi-copy gene families, leading to copy number instability through unequal crossing over, recombination, and transcription-associated breaks [10–13]. In model organisms, rDNA copy number variation influences traits from lifespan in yeast to metabolic programming and body size in mice [14–16]. Clinical observations reinforce these findings: ribosomopathies, genetic disorders of ribosome biogenesis, cause anemia, growth defects, and metabolic dysfunction [17, 18]. Together, these observations suggest that variation in rDNA copy number may have unrecognized consequences for human health.

Historically, the repetitive nature of rDNA has excluded it from most large-scale genetic analyses. However, advances in whole-genome sequencing and extensively phenotyped cohorts now enable systematic investigation [19–21]. Initial population-scale studies of the 45S array demonstrated high heritability and associations with hematological profiles and renal function markers, though limited sample sizes constrained the detection of disease associations [22]. The 5S array has not been examined at population scale, and no study has assessed whether the two arrays have independent or coordinated effects on human phenotypes.

Here, we leverage 490,383 UK Biobank participants to perform the largest analysis of 45S rDNA copy number to date and the first population-scale characterization of 5S. We trace population-level associations through physiological traits, plasma proteomics from 54,219 UK Biobank participants, and tissue-specific gene expression across 57 tissues from 948 GTEx donors, then examine functional consequences in primary human pancreatic islets. These arrays supply essential components to ribosomes but behave as independent genetic factors with divergent consequences: 45S with metabolic disease, adiposity, hematological signatures, and altered insulin secretion in primary human pancreatic islets organoids; 5S with proportional organismal scaling, including increased lean mass and organ volumes, alongside reduced insulin response to glucose stimulation. Across scales, these molecular signatures align with population-level phenotypes, establishing that natural copy number variation in the two rDNA arrays has divergent consequences for human health and disease.

## 3 Results

### 3.1 Quantification of 45S and 5S rDNA CN across the UK Biobank reveals extensive population variation

To characterize rDNA variation at population scale, we quantified 45S and 5S copy number in 490,383 UK Biobank participants using whole-genome sequencing data (**Figure 1A**). Our approach leveraged the Telomere-to-Telomere consortium’s complete rDNA consensus sequences [23] to map rDNA-containing coordinates on the GRCh38 reference genome, followed by GC-corrected read depth normalization and batch effect correction (see **Methods** and (**Supplementary Figure 2**). This represents the largest analysis of 45S rDNA to date and the first population-scale characterization of 5S, enabling detection of disease associations not observed in previous studies of smaller cohorts [22]. To trace the biological consequences of rDNA variation from population-level associations to cellular function, we integrated these copy number estimates with plasma proteomics, tissue-specific gene expression, and primary human islet phenotypes (**Figure 1B**).

**Figure 1:**
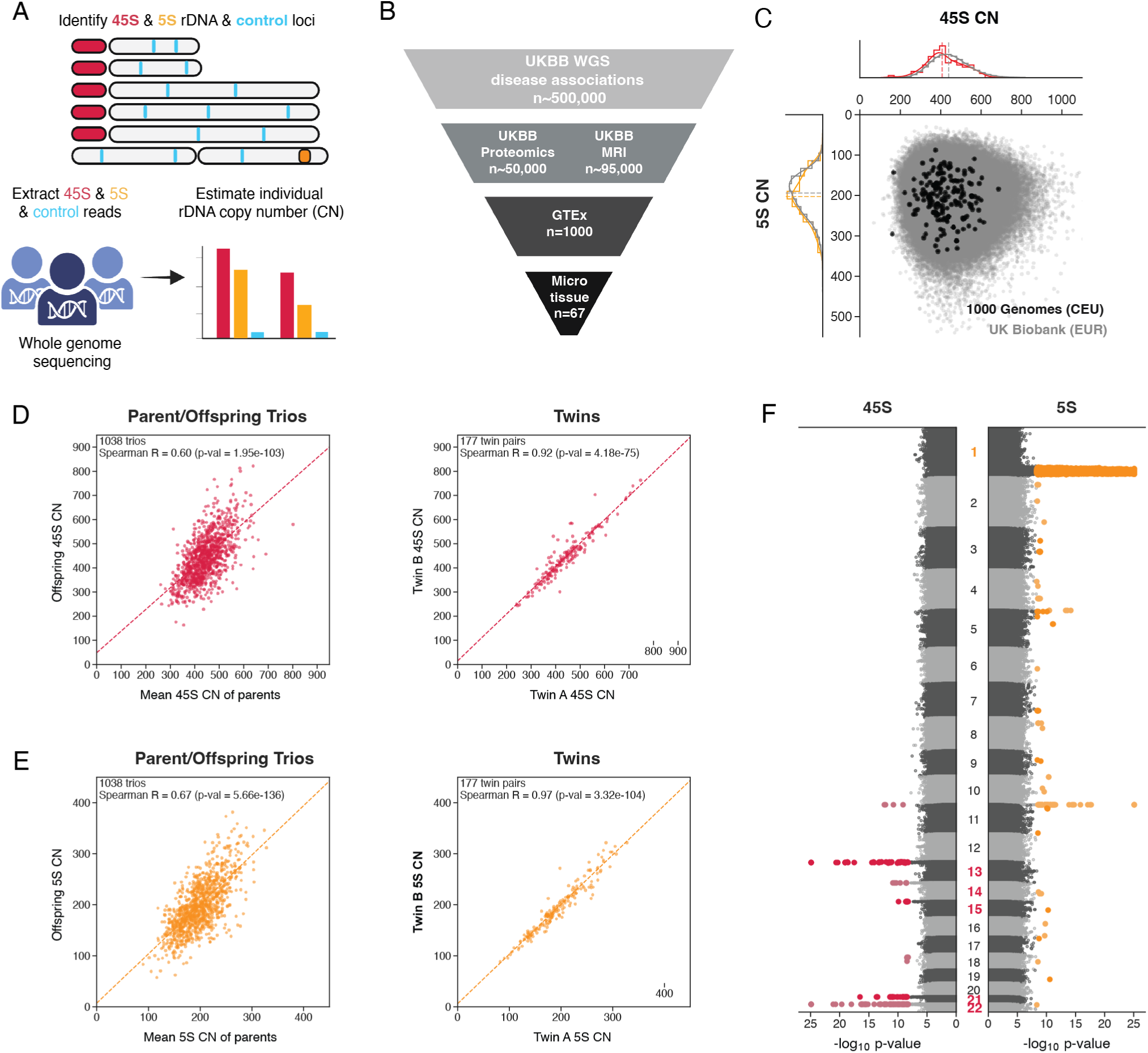
rDNA 5S and 45S copy numbers are variable and heritable in the UK Biobank population. **A**: Organization of human rDNA arrays and the workflow to quantify rDNA copy number (CN). **B**: Multi-scale analysis framework integrating rDNA copy number with disease associations, plasma proteomics, tissue-specific gene expression, and primary islet function. **C**: CN variation of 45S and 5S rDNA in the UK Biobank (grey) and the 1000 Genomes cohort (black). **D**: Concordance of 45S CN among parent-offspring trios (left) and monozygotic twins (right) within the UK Biobank. **E**: Concordance of 5S CN among parent-offspring trios (left) and monozygotic twins (right) within the UK Biobank. **F**: Manhattan plot showing genetic variants associated with 45S CN (red) and 5S CN (orange). Chromosome numbers are colored in red for acrocentrics containing 45S and orange for Chromosome 1 containing the 5S array.

The resulting copy number distributions revealed substantial inter-individual variation, with the average diploid genome harboring 450 45S copies (std. dev. = 97) and 198 5S copies (std. dev. = 50) (**Figure 1C**). These estimates showed high concordance with published values from an alternative smaller cohort [24] (1000 Genomes; Spearman *ρ* = 0.93 for 45S and *ρ* = 0.96 for 5S; **Supplementary Figure 3**) [25], and distributions were consistent across genetic ancestry groups (**Supplementary Figure 4**).

Despite both arrays encoding essential ribosomal RNA components, 45S and 5S copy numbers showed negligible correlation across the cohort (Spearman *ρ* = 0.017, *r*^2^ *<* 0.001; **Figure 1C**), consistent with previous observations in smaller cohorts [25]. The statistically significant p-value reflects sample size rather than biological coordination. This independence indicates that the two arrays, though functionally linked in ribosome assembly, vary as genetically distinct entities. To confirm our measurements reflect stable germline variation, we examined concordance among relatives: both arrays showed high concordance in parent-offspring trios (Spearman *ρ* = 0.60 and 0.67 for 45S and 5S, respectively) and monozygotic twins (*ρ* = 0.92 and 0.97; **Figure 1D-E**). Minimal correlation with age (*ρ* = *−*0.008 for 45S, *ρ* = 0.005 for 5S) indicates that these estimates reflect germline copy number rather than age-related somatic changes.

Given the high concordance of rDNA copy number within trios, we performed genome-wide association analysis to identify genetic loci influencing each array. This revealed multiple genome-wide significant associations for both 45S and 5S (**Figure 1F**), in contrast to a previous study of approximately 220,000 individuals that had not identified any significant loci [22]. The strongest signals for each array mapped to cis-acting loci on the rDNA-containing chromosomes themselves: the acrocentric chromosomes for 45S and chromosome 1 for 5S, consistent with findings of active recombination on rDNA arrays and specifically within recently described pseudohomologous regions. [12, 13, 26]. Notably, both arrays showed significant associations on chromosome 11, suggesting shared trans-regulatory mechanisms that warrant further investigation. The distinct linkage architectures revealed by GWAS, combined with the absence of copy number correlation, establish that 45S and 5S vary as independent genetic factors despite their coordinated function in ribosome biogenesis.

### 3.2 45S but not 5S rDNA copy number is associated with diverse human diseases

To investigate the phenotypic consequences of rDNA variation, we performed phenome-wide association studies between rDNA CN and disease outcomes in UK Biobank participants of European ancestry. This analysis revealed extensive associations between 45S copy number and human disease (**Figure 2A**, upper panel). Higher 45S copy number associated with increased risk across multiple organ systems, with the strongest signals in metabolic, cardiovascular, and renal diseases. These included type 2 diabetes (*P* = 2.07 *×* 10*^−^*^5^), acute myocardial infarction (*P* = 9.8 *×* 10*^−^*^5^), and chronic kidney disease (*P* = 2.05 *×* 10*^−^*^8^). Diseases of the respiratory system also showed associations, including chronic obstructive pulmonary disease (*P* = 3.04 *×* 10*^−^*^5^) and asthma (*P* = 8.9 *×* 10*^−^*^6^). This broad pattern of disease associations was unique to 45S. When we performed the same analysis for 5S copy number, we found no significant disease associations despite equivalent statistical power (**Figure 2A**, right panel).

**Figure 2:**
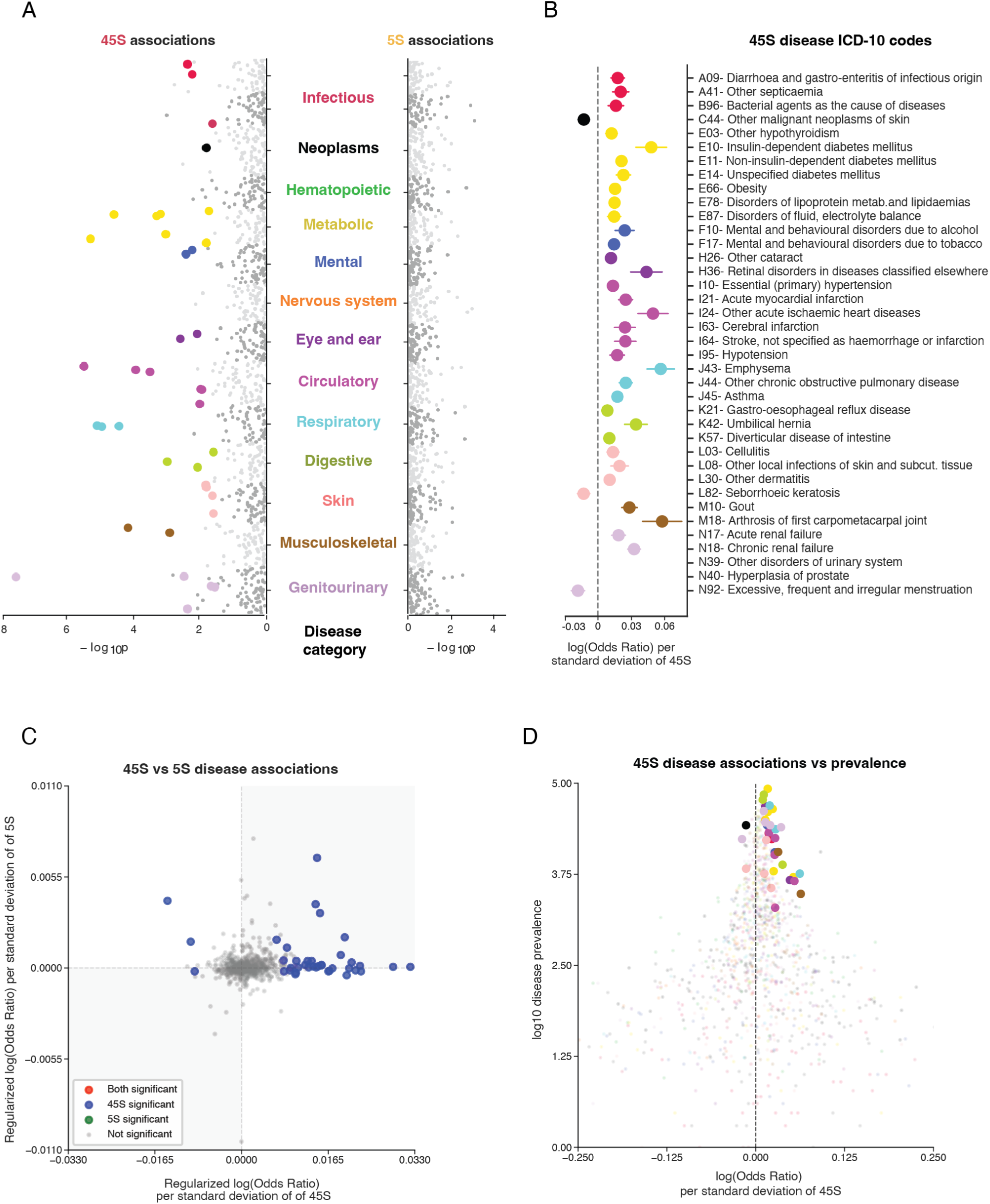
Unlike 5S, 45S rDNA copy number is associated with human disease. **A**: Disease associations for 5S (left) and 45S (right) rDNA CN. Significant associations are shown as large points, the colors represent disease groups as defined by ICD-10-CM. **B**: Log odds ratios for significant disease associations in A for 45S. **C**: Correlation of disease log odds ratio (shrinkage estimate) between 45S and 5S. **D**:Comparison of disease log odds ratio for 45S vs. the disease prevalence.

To contextualize the magnitude of 45S associations, we examined effect sizes per standard deviation of copy number (**Figure 2B**). With a population standard deviation of approximately 95 copies, individuals at the 16th percentile versus the 84th percentile differ by roughly 190 copies. Using the association of type 2 diabetes as example, this translates to an approximately 12% difference in disease risk. For individuals at the extremes of the distribution (5th vs 95th percentile, approximately 400 copies difference), odds ratios approach 1.2 for several conditions including ischemic heart disease, retinal disorders, and emphysema.

To confirm this divergence reflected truly independent effects rather than attenuated 5S effects failing to reach significance, we examined the correlation of effect sizes across all diseases tested. The absence of correlation (**Figure 2C**) demonstrates that these arrays have fundamentally distinct relationships with disease risk.

The strength of disease associations depends on disease prevalence in this generally healthy cohort (**Figure 2D**). Common metabolic and cardiovascular conditions show robust associations, while rarer outcomes including neurodegenerative diseases and mortality do not reach significance (**Supplementary** Figures 6**, 7**). These patterns reflect a fundamental limitation of disease-based analyses: binary outcomes in a relatively healthy cohort constrain detection to common conditions. To characterize the full spectrum of physiological effects beyond diseases, we next examined continuous traits and molecular phenotypes measured across the entire population.

### 3.3 Quantitative traits reveal divergent physiological signatures of 5S and 45S copy number variation

Having established that disease associations are limited to 45S, we examined continuous traits across the UK Biobank to characterize the full physiological spectrum of rDNA variation. The two arrays showed divergent signatures across hematological, anthropometric, and metabolic phenotypes (**Figure 3A**).

**Figure 3:**
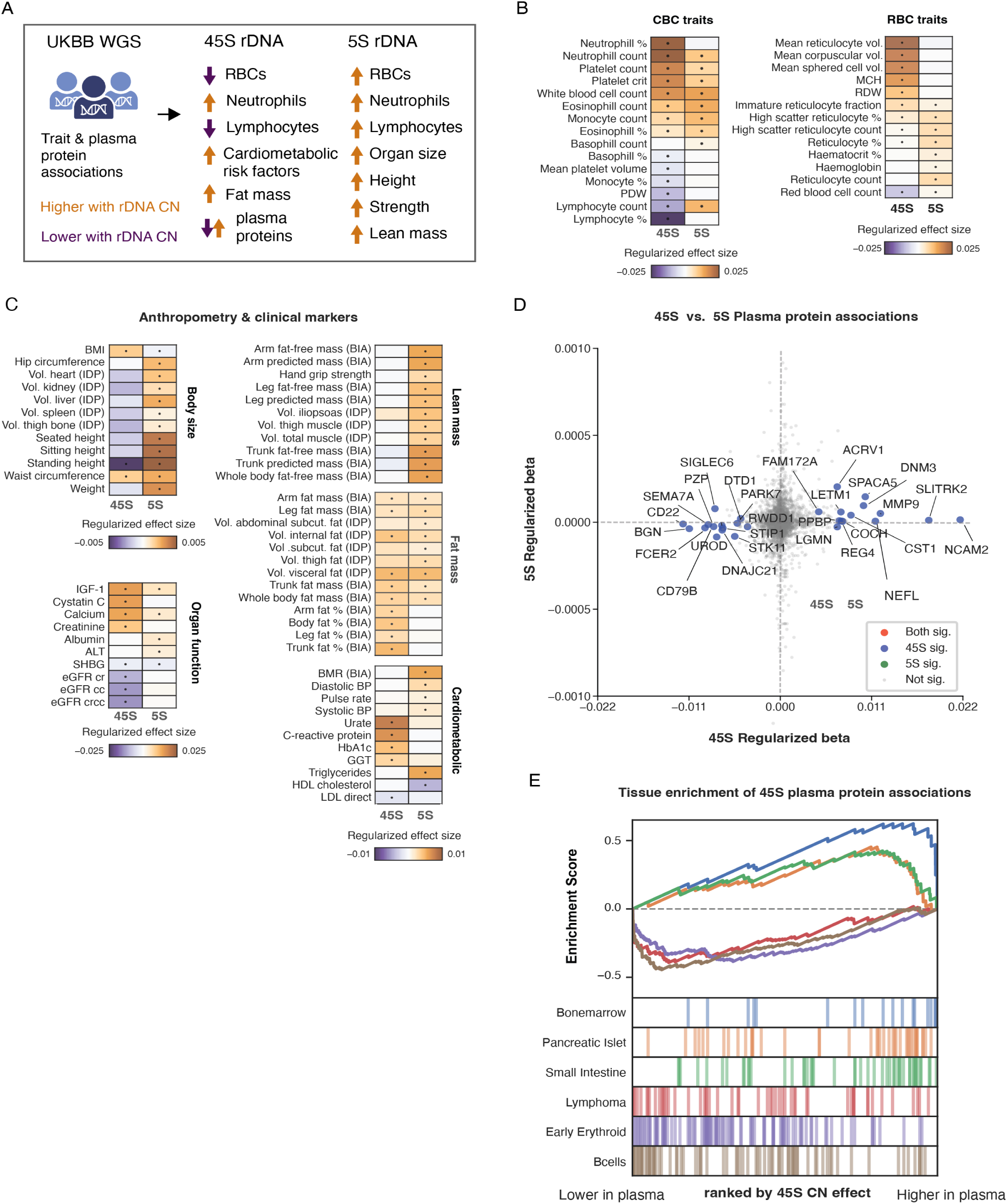
Divergent physiological signatures of 45S and 5S rDNA copy number variation. **A**: Summary of divergent phenotypic associations for 45S and 5S rDNA CN across traits and plasma proteins. **B**: Associations of rDNA 45S and 5S CN with complete blood count (CBC) and red blood cell (RBC) parameters. Black dots indicate significance. **C**: Anthropometric traits and clinical biomarkers. **BIA** = Bioelectrical Impedance Analysis. **IDP** = Image Derived Phenotype. **D**: Comparison of 45S and 5S effect sizes on plasma protein abundance. **E**: Gene Set Enrichment Analysis (GSEA) of plasma proteins ranked by 45S CN effect size using Human Gene Atlas tissue profiles.

Complete blood count parameters revealed distinct hematopoietic effects (**Figure 3B**). Higher 5S copy number correlated with increases across all hematopoietic lineages: elevated absolute counts of neutrophils, lymphocytes, monocytes, and eosinophils, while percentages of individual cell types remained unchanged. This pattern indicates balanced hematopoietic expansion without altered cellular ratios. In contrast, 45S showed selective changes consistent with inflammation and erythrocyte dysfunction. Higher 45S copy number associated with increased neutrophil percentage while decreasing lymphocyte percentage and platelet counts, an inflammatory signature consistent with previous causal analyses [22]. A distinct pattern emerged in erythrocyte parameters: higher 45S corresponded to decreased red blood cell counts, increased mean corpuscular volume, elevated mean corpuscular hemoglobin, and higher red cell distribution width. This macrocytic pattern is reminiscent of ribosomopathies, though concurrent neutrophilia distinguishes natural rDNA variation from ribosome biogenesis mutations [17, 18, 27].

Body composition showed consistent divergence between 45S and 5S copy number across three orthogonal measurement modalities: standard anthropometry, and bioelectric impedance analysis (BIA) from the whole cohort, and MRI-based image-derived phenotypes (IDPs) from 95,665 participants (**Figure 3C**; Methods). Higher 45S copy number correlated specifically with adiposity: while both arrays associated with increased fat mass, only 45S increased body fat percentage across multiple depots. This selective increase in fat percentage rather than absolute mass suggests adipose accumulation relative to lean tissue, consistent with the metabolic disease associations and previous reports of association with increased BMI [15]. Higher 5S copy number instead associated with increased body size and lean mass, including fat-free mass and standing height. This pattern extended to the organ level, where 5S copy number correlated with increased volumes of heart, kidney, liver, spleen, and thigh bone as measured by MRI (**Figure 3C**, Body size). The concordance between classic anthropometry, BIA, and MRI-based IDPs across independent measurement modalities suggests distinct underlying biology rather than measurement artifact. Clinical chemistry biomarkers reinforced these divergent compositional profiles (**Figure 3C**). Higher 45S correlated with markers of metabolic syndrome: elevated HbA1c, C-reactive protein, urate, creatinine, cystatin C, and gamma-glutamyltransferase, alongside reduced eGFR [28,29]. This biochemical profile aligns with the observed associations with type 2 diabetes, cardiovascular disease, and chronic kidney disease. Higher 5S showed a distinct anabolic profile with elevated albumin and triglycerides. These biochemical signatures establish that 45S variation correlates with inflammatory dysfunction and metabolic syndrome, while 5S variation associates with increased metabolic capacity. To examine the molecular basis of these associations, we next analyzed the plasma proteome measured in a subset of the UK Biobank cohort.

### 3.4 Plasma proteomics reveals 45S-specific effects on secretory tissues

To examine the molecular basis of 45S disease associations, we analyzed plasma proteomics from 54,219 UK Biobank participants, measuring approximately 3,000 proteins via antibody-based assays [30] (**Figure 3D**). Kidney dysfunction substantially influences circulating protein profiles, and 45S copy number associated with chronic kidney disease; to ensure associations were not secondary to renal function, we included eGFR as a covariate and found that results persisted with similar magnitude and direction (**Supplementary Figure 9**).

We identified proteins significantly associated with 45S CN at 10% FDR. The strongest positive associations included neural damage markers (NEFL, NCAM2, SLITRK2), inflammatory mediators (MMP9), and tissue remodeling proteins. The prominence of NEFL, a marker of neuronal distress that typically appears in circulation only during neuronal damage [31], suggests neurological stress in individuals with higher 45S CN. This finding is notable given the absence of neurodegenerative disease associations, likely reflecting low disease prevalence rather than absent biological effects (**Figure 2C**). Proteins showing negative associations were dominated by B-cell markers (CD22, CD79B, FCER2, SIGLEC6), consistent with the reduced lymphocyte percentages observed in blood counts (**Figure 3B**). 5S copy number showed no significant associations with the plasma proteome despite effects on blood composition and body mass, reinforcing the distinct biology of these arrays.

To identify the tissue origins of 45S-associated proteins, we performed Gene Set Enrichment Analysis using Human Gene Atlas tissue profiles [32] (**Figure 3E**). This revealed enrichment for proteins specifically expressed in bone marrow, pancreatic islets, and small intestine, with low 45S copy number associating with erythroid and B-cell markers. Analysis using single-cell-derived gene sets further refined these signals, revealing enrichment for secretory cell types including goblet cells, paneth cells, and acinar cells (**Supplementary Figure 8**) [33]. These cell types share extreme translational demands: goblet cells dedicate up to 20% of cellular volume to mucin production and paneth cells continuously secrete antimicrobial peptides, consistent with the small intestine enrichment in tissue-level analysis; acinar cells produce massive quantities of digestive enzymes, aligning with the pancreatic islet signal [34, 35]. The convergence on high-output secretory cells suggests that 45S copy number variation most strongly affects tissues operating at the limits of protein synthesis capacity.

### 3.5 Tissue-specific gene expression reveals distinct molecular programs for 45S and 5S copy number variation

To examine whether the divergent phenotypic associations of 45S and 5S reflect distinct molecular programs, we analyzed gene expression across 57 tissues in the Genotype-Tissue Expression (GTEx) cohort using matched whole genome sequencing and RNA sequencing data from 948 postmortem donors (**Figure 4A**) [36]. We quantified rDNA copy number from GTEx whole genome sequencing data and observed similar distributions and array independence as in the UK Biobank (**Supplementary Figure 13**). We then tested the effect of both arrays on gene expression in each tissue independently, controlling for age, sex, and technical covariates, and performed pathway enrichment analysis on the resulting data (**Figure 4B**). Enrichment was assessed agnostic to effect direction, as the same pathway may be upregulated in one tissue and downregulated in another depending on cellular context; the goal was to identify which biological processes are responsive to rDNA variation regardless of the direction of that response.

**Figure 4:**
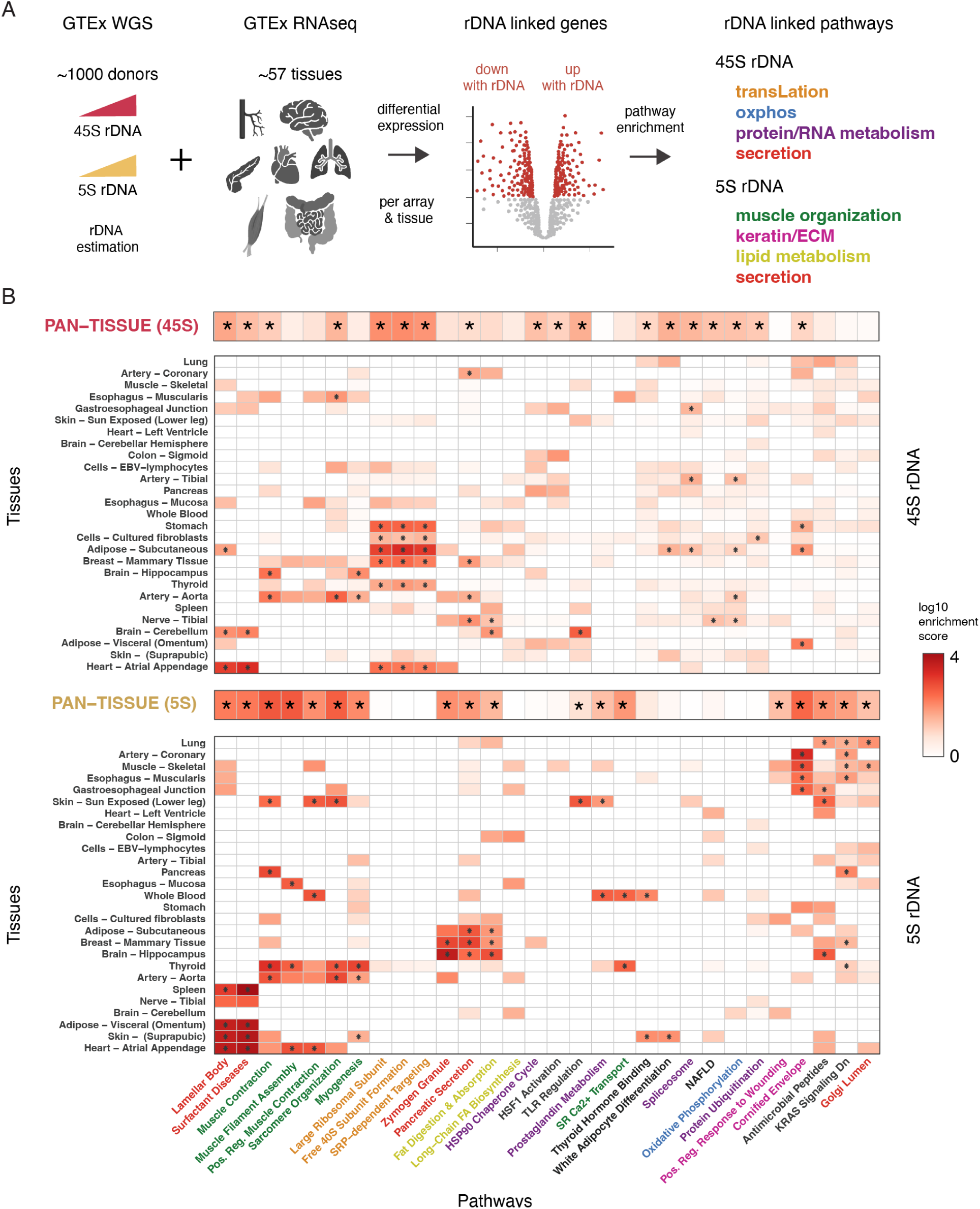
Tissue-specific gene expression reveals distinct molecular programs for 45S and 5S rDNA copy number variation. **A**: Schema for GTEx analysis workflow integrating whole genome sequencing for rDNA copy number estimation with RNA sequencing across 57 tissues from 948 donors. **B**: Pathway enrichment heatmaps for 45S (top) and 5S (bottom) copy number associations. Columns represent pathways significant in pan-tissue analysis (stars); rows represent tissues with significant gene associations for both arrays. Color intensity indicates enrichment score.

45S copy number showed broad pathway enrichment across tissues, with pan-tissue significance for translation machinery (ribosomal subunit formation, SRP-dependent targeting), cellular stress responses (HSF1 activation, HSP90 chaperones), and secretory processes (pancreatic secretion, fat digestion and absorption). Particularly strong enrichment emerged in heart, thyroid, breast, and stomach. These pathways align with the canonical functions of 45S-encoded ribosomal RNAs and provide molecular context for the disease associations: elevated translational and secretory burden may underlie the metabolic dysfunction observed in individuals with higher 45S copy number.

5S copy number yielded fewer significant genes overall (**Supplementary Figure 11**), and the pathways that reached significance in pan-tissue analysis were dominated by muscle biology: muscle contraction, myofibril assembly, sarcomere organization, and myogenesis. These muscle-related pathways showed enrichment across diverse tissues rather than exclusively in muscle, suggesting systemic effects on myogenic gene programs. Additional 5S-associated pathways included lipid metabolism terms such as fat digestion and absorption in subcutaneous adipose. This molecular signature aligns with the UK Biobank anthropometric findings, where 5S copy number correlated with increased lean mass, grip strength, and proportional organismal scaling.

While significance-based pathway analysis highlighted divergent enrichment patterns between arrays, direct examination of effect sizes revealed a more nuanced picture. Both 45S and 5S copy number showed concordant positive associations with ribosomal protein gene expression across most tissues (**Supplementary Figure 12**). However, 45S effects were larger and more consistent across tissues, producing robust statistical significance after multiple testing correction, whereas 5S effects were more modest and variable, falling below detection thresholds in most individual tissues. This suggests that both arrays influence translational capacity in the same direction, but 45S exerts stronger and more uniform effects. The distinct pathway signatures in Figure 4 therefore reflect differences in effect magnitude rather than opposing biological programs.

The convergence of transcriptomic signatures with population-level phenotypes across independent cohorts strengthens the case that natural rDNA copy number variation has systematic effects on gene expression programs. These molecular distinctions provide context for the divergent anthropometric associations: 45S variation, with its stronger translational effects, may primarily affect metabolic and secretory tissues, whereas 5S variation shapes lean mass and musculoskeletal phenotypes through its influence on myogenic gene programs.

### 3.6 rDNA copy number influences insulin secretion in human pancreatic islets

To examine whether rDNA copy number affects cellular function in a disease-relevant tissue that appeared in several of our prior analyses, we studied the effect in primary human pancreatic islet microtissues from 67 donors (47 male, 20 female). We performed whole genome sequencing to quantify rDNA copy number and correlated these measurements with standardized glucose-stimulated insulin secretion (GSIS) assays (**Figure 5A**). The rDNA copy number distributions matched those observed in UK Biobank, confirming technical consistency (**Figure 5B**).

**Figure 5:**
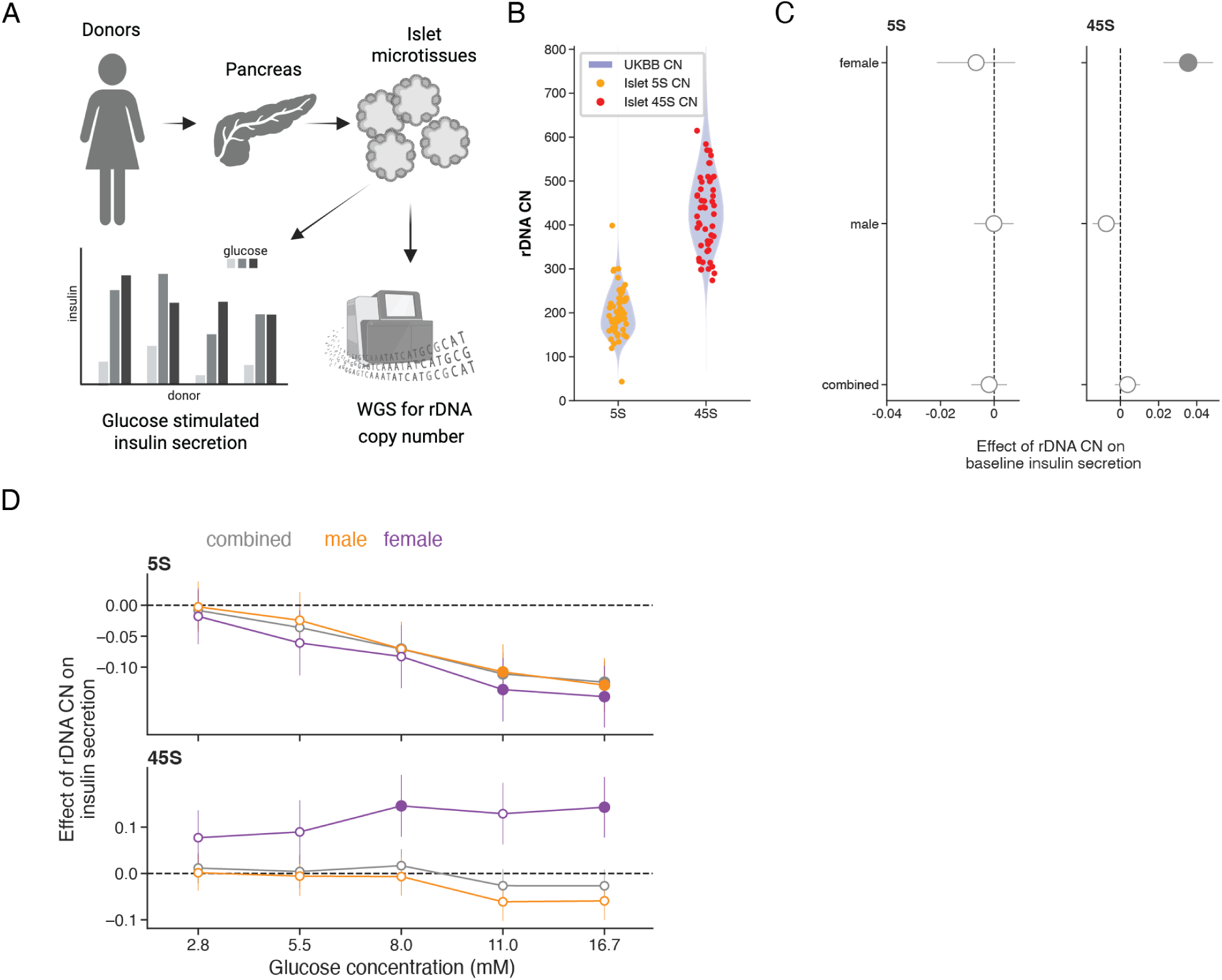
rDNA copy number modulates insulin secretion in pancreatic islets. **A**: Workflow to measure rDNA copy number and glucose-stimulated insulin secretion (GSIS) in pancreatic islets from human donors. **B**: rDNA copy number estimates from donor-derived pancreatic islets compared with estimates derived from whole blood in UK Biobank participants. **C**: Sex-dependent effect of 5S and 45S rDNA copy number on baseline insulin secretion in human pancreatic islets. **D**: Sex-dependent effect of 5S and 45S rDNA copy number on insulin secretion as a function of glucose concentration.

Using a linear mixed effects model incorporating all glucose concentrations, biological replicates, and donor characteristics, we identified divergent associations between the two rDNA arrays and insulin secretion. Higher 45S copy number associated with increased basal insulin secretion and enhanced glucose-stimulated insulin release specifically in fe-male donors, while males showed no association (**Figure 5C-D**). In contrast, higher 5S copy number associated with reduced insulin secretion under high glucose conditions in both sexes (**Figure 5C**). This sex specificity in islets contrasts with disease associations, which showed no sex-dependent effects in the UK Biobank (**Supplementary Figure 5**).

The opposing effects of 45S and 5S on insulin secretion may align with the secretion-related pathway enrichment observed for both arrays in GTEx, though the mechanisms underlying their divergent functional consequences remain to be determined. While these in vitro findings cannot establish causal links to diabetes risk or explain the disease associations observed at the population level, they nevertheless demonstrate direct functional effects of rDNA copy number variation on insulin secretion for the first time.

## 4 Discussion

The two human ribosomal DNA arrays, despite encoding essential components of the same molecular machine, vary independently and associate with distinct consequences for human health. Higher 45S copy number correlates with metabolic disease, adiposity, and inflammatory markers, while 5S copy number associates with proportional organismal scaling without disease risk.

The convergence of findings across independent cohorts and biological scales strengthens the case that these associations reflect genuine biology rather than technical artifacts or isolated correlations. Population-level disease associations in nearly 500,000 UK Biobank participants were recapitulated in plasma proteomics from 54,000 individuals, tissue-specific gene expression across 57 tissues from 948 GTEx donors, and functional assays in primary human pancreatic islets. At each scale, 45S and 5S showed distinct signatures consistent with their population-level phenotypes: 45S with secretory and translational stress, 5S with myogenic programs and metabolic capacity.

The molecular signatures associated with 45S variation suggest a coherent pathophysiological model. Plasma proteomics revealed elevated proteins from high-output secretory cell types including pancreatic islets, small intestine, and bone marrow. GTEx analysis showed pan-tissue enrichment for translation machinery, stress responses, and secretory pathways. In islets, higher 45S copy number correlated with enhanced glucose-stimulated insulin secretion. Together, these findings suggest that 45S variation modulates secretory burden, with tissues operating at the limits of protein synthesis capacity affected the most. The hematological signatures reminiscent of ribosomopathies, including macrocytic indices and altered erythropoiesis, further support a model in which natural 45S variation produces subclinical perturbations in ribosome-dependent cellular functions [17,18]. This interpretation aligns with the nucleolar localization of 45S transcription, which couples rRNA synthesis to growth signaling and metabolic stress sensors [37, 38].

The absence of disease associations for 5S variation despite substantial physiological effects presents an intriguing contrast. 5S copy number associates with coordinated organismal scaling: increased lean mass, organ volumes, grip strength, and basal metabolic rate. GTEx analysis revealed enrichment for myogenic gene programs across diverse tissues, providing molecular validation of the anthropometric findings. Several features of 5S biology may contribute to this divergence from 45S. 5S rRNA is transcribed by RNA polymerase III in the nucleoplasm, spatially and transcriptionally separate from nucleolar 45S biogenesis [39]. Additionally, 5S rRNA participates in extra-ribosomal functions through the RPL5-RPL11-MDM2-p53 checkpoint [9], raising the possibility that cells buffer 5S variation through regulatory pathways rather than pathological stress responses. Whether these mechanisms explain the absence of disease associations requires direct experimental investigation.

Notably, both arrays showed enrichment for secretion-related pathways in GTEx, yet their effects on insulin secretion were opposing. The shared involvement in secretory biology, with different tissue distributions but overlapping pathway terms, suggests both arrays influence aspects of protein production and trafficking, though by mechanisms that remain to be defined.

Several limitations warrant consideration. The sex-specific effects observed in pancreatic islets, where 45S associations with insulin secretion were restricted to female donors, contrast with the absence of sex interactions in UK Biobank disease associations (**Supplementary Figure 5**). This discrepancy may reflect cohort-specific features, statistical power, or genuine biological complexity that our current data cannot resolve. More fundamentally, while the multi-scale convergence provides strong correlative evidence, our findings do not establish causal relationships between rDNA copy number and disease risk. The repetitive, essential nature of rDNA arrays makes them refractory to standard genetic perturbation approaches, and the mechanisms by which copy number variation influences cellular physiology remain to be defined.

Beyond copy number, rRNA sequence variation represents an additional axis of ribosomal diversity with phenotypic consequences. Complementary analysis of rRNA sequence variants in the UK Biobank revealed that heritable variants cluster within expansion segments of the 28S rRNA, with distinct regions associating with different trait categories: es15l with adiposity, es39l with body dimensions, and es27l with blood phenotypes and disease susceptibility [40, 41]. Notably, both copy number and sequence variation associate with overlapping phenotypic domains including adiposity and metabolic traits, yet operate largely independently of one another. Together, these findings establish that rDNA harbors multiple dimensions of functional variation that have been systematically excluded from genome-wide analyses.

Together, our findings position rDNA copy number variation as an underexplored axis of human phenotypic variation. The effect sizes for disease associations are comparable to established genetic risk factors, and the convergence across molecular and physiological scales suggests that rDNA variation has systematic consequences for human biology. Understanding how cells balance the independent variation of these two essential arrays, and why imbalance produces disease risk for one but not the other, may reveal fundamental principles of ribosome homeostasis with implications for metabolic disease.

## 5 Methods

### 5.1 Human subjects

This research was conducted using the UK Biobank Resource under applications 18448 and 44584. UK Biobank received ethical approval from the NHS National Research Ethics Service (ref: 11/NW/0382); all participants provided written informed consent. GTEx data were obtained under dbGaP accession phs000424; the GTEx project was approved by institutional review boards at all participating sites and all donors or their families provided informed consent. Primary human pancreatic islet microtissues were obtained from InSphero AG (Schlieren, Switzerland), derived from anonymized deceased organ donors with consent from next of kin obtained by the upstream tissue procurement organizations. No tissues were procured from prisoners. Donor characteristics are summarized in Supplementary Table X; race/ethnicity classifications were provided by the vendor and were not used in statistical analyses.

### 5.2 Identifying ribosomal DNA reads from WGS data

A key challenge in analyzing genetic variation at highly repetitive genomic loci with short-read WGS data is determining their location on the GRCh38 reference build of the human genome. To resolve this, we started with the annotated 5S sequences (on chr1) and 45S sequences (on the acrocentric chromosomes chr13, chr14, chr15, chr21, and chr22) from the most complete reference human genome published by the Telomere-to-Telomere consortium [23]. We simulated 150bp paired-end reads from these annotated sequences and mapped them to the reference human genome GRCh38. This is the same reference to which the WGS data were mapped, by both the 1000 Genomes project [24] and by UK Biobank [42]. We identified three loci on chr1 in the GRCh38 build to which simulated 5S reads mapped, and five loci across contigs `chr21`, `chr22_KI270733v1_random`, and `chrUn_GL000220v1` in GRCh38 to which simulated 45S reads mapped.

For each sample, we filtered reads mapping to the rDNA loci (henceforth, referred to as rDNA reads) from the associated CRAM file. Across all samples, we observed a roughly equal split between properly paired and improperly paired rDNA reads, which is consistent with our expectation given that the tandem 5S and 45S arrays are collapsed within the GRCh38 genome build. Finally, at each position within the rDNA loci and for each choice of fragment (read-pair) length, we enumerated the number of fragments where the 5’ end of the fragment aligned to that position. This resulted in sparse LxM count matrices, one each for the 5S and 45S subunit, where L is the combined length of the rDNA loci and M is the maximum fragment length (set to 1000 bp).

### 5.3 Estimating copy number of the 5S and 45S subunits

The total number of mapped rDNA fragments obtained from these count matrices is proportional to the number of copies of the rDNA subunits. In addition, sequencing depth, fragment-size distribution, and GC-biased PCR amplification also contribute to variation in the total rDNA read count across samples. We correct for these sources of variation by modeling the fragment counts at each position using a Poisson model whose rate is a function of the GC-content of the fragment sequence [43]. We used read-pairs mapped to a fixed set of 1000 control loci, matched in length to the rDNA loci, to estimate the rate of observing fragments as a function of its GC-content (allowing for a diploid genome). This rate is estimated for each sample separately, and, thus, is implicitly a function of sequencing depth and the median fragment-size. We used these in-sample *fragment rates* to compute the expected number of fragments mapping to the rDNA locus. Finally, we estimated the 45S (similarly, 5S) copy number as the ratio of the observed total 45S (and 5S) fragments to this expected number of 45S (and 5S) fragments. The 45S estimates were multiplied by 6.29 and the 5S estimates were multiplied by 18.41, to account for the difference in length between a single unit of each array (43Kb for 45S and 2.2Kb for 5S) and their corresponding loci on the GRCh38 build (270Kb for 45S and 41Kb for 5S).

### 5.4 Correcting for batch effects from population-scale sequenc-ing

Whole genome sequencing of UK Biobank participants were conducted in two phases (Vanguard phase and Main phase), across multiple sequencing and bioinformatics providers. In addition to CRAM files, uniformly processed using DRAGEN v3.7.8, the UK Biobank has provided several quality control metrics associated with generating and processing these data (Category 187 on UK Biobank showcase). We explored the impact of those QC metrics available for the entire cohort on our estimates of 45S and 5S CN. While most QC metrics had a significant association (*p <* 0.01) with our CN estimates, the sequencing provider (Field 32051) had the strongest association on 45S CN (**Supplementary Figure 2**. We corrected our estimates for rDNA CN by regressing out the effects of significantly associated QC metrics.

### 5.5 Determining genetically related participants in UK Biobank

The UK Biobank provides genetic relatedness information for 107,162 pairs of participants, covering a total of 147,753 participants. Participant pairs with kinship *>* 0.35 and matched birth years (Field 34) were considered monozygotic twins. Participant pairs with kinship between 0.17 and 0.35, and birth years differing by more than 18 years were considered parent-offspring pairs. We used these criteria to identify 172 monozygotic twins and 1012 trios with WGS data in the UK Biobank cohort.

### 5.6 Genomewide association study of rDNA variation

We tested for association between 45S (and 5S) CN and all genotyped and imputed genetic variants (Field 21008) with INFO score greater than 0.7, using REGENIE v3.4.1 [44]. We restricted our analysis to 420,705 individuals of EUR ancestry (Field 30079), and controlled for sex (Field 22001), recruitment center (Field 54), genotyping array (Field 22000), and 20 genetic principal components [45].

### 5.7 Testing effects of rDNA variation on complex traits in UK Biobank

We tested for association between 45S (and 5S) CN and a range of molecular, cellular, and organismal traits such as anthropometric traits, plasma protein abundances, levels of clinical biomarkers, blood cell counts, and disease incidence. All quantitative traits were rank-inverse normal transformed before testing for association. We restricted our discovery association analyses to individuals in the EUR ancestry group to maximize power. For each trait, we tested for the additive effect and the sex-interaction effect of rank-inverse transformed CN on the trait, controlling for the effects of recruiting center (Field 54) and principal components (estimated by the Pan-UK Biobank project) [45].

#### 5.7.1 Disease incidence

The first reported occurrence of each disease ICD10 code for each participant was obtained from Fields 130000 to 132606. We processed these first occurrences into binary disease incidence measures for 1,257 3-digit ICD10 codes across the complete UK Biobank cohort. We excluded codes with fewer than 10 diagnosed cases within the EUR cohort in UK Biobank, and tested for association between CN and disease incidence for the remaining 1,098 ICD10 codes using logistic regression.

#### 5.7.2 Anthropometric traits

This category of traits included standing height (Field 50), seated height (Field 51), sitting height (Field 20015), hip circumference (Field 49), waist circumference (Field 48), grip strength (Fields 46 and 47), weight (Field 21002), BMI (Field 21001), systolic and diastolic blood pressure (Fields 4079 and 4080; average of repeat measurements), pulse rate (Field 102; average of repeat measurements), and traits derived from whole body bio-impedance measurements (Category 100009).

#### 5.7.3 MRI-derived traits

This category of traits included volume measurements of organs and specific depots of fat derived from deep-learning based segmentation of whole body magnetic resonance images [46–48]. To ensure high-quality segmentations with full anatomical coverage, thresholding criteria were applied. Ranges were defined for total muscle (5–60 L) and thigh intramuscular adipose tissue (1–15 L). These upper volume limits were defined following visual inspection of extreme values to exclude spurious segmentations caused by leakage into adjacent structures. Minimum volumes were set for kidneys (30 mL / kidney) to exclude partial or failed segmentations, while inter-vertebral discs were capped at 1 L. Vessel cross-sectional areas (CSAs) were filtered to exclude irregular roundness (*<* 0.7), aortic arch CSA (*>* 1600 mm^2^). The superior vena cava was informed by visual inspection of extreme values (99.99^th^ percentile of the CSA distribution) with exclusions applied on the cases showing clear segmentation errors. Zero-volume major organs (liver, spleen, pancreas, lungs, heart) and sparse 2D slice segmentations (*<* 10 voxels) were removed. These filters removed between an average of 0.4% of values per imaging-derived phenotype (range: 0.01% *−* 10.9%).

#### 5.7.4 Clinical biomarkers

This category of traits included key blood biochemistry markers (Category 17518) and es-timated Glomerular Filteration Rate (eGFR). GFR estimates were derived from measurements of Creatinine (Field 30700) and Cystatin C (Field 30720), using formulae adopted by the National Kidney Foundation (https://www.kidney.org/professionals/gfrcalculator).

#### 5.7.5 Plasma proteome

This category of traits included plasma protein abundances, as quantified by normalized protein expression values measured using the Olink platform. Plasma abundance was measured for 2942 proteins across 53,058 participants (Field 30900). For this category of traits, we controlled for the following additional covariates: hour of blood collection, month of blood collection, number of years the sample was stored, fasting time at blood collection, plate used for sample run (Field 30901) and the first two principal components for each panel of proteins. Gene Set Enrichment Analysis (GSEA) was conducted using the GSEApy Python package. Proteins were pre-ranked based on the product of the ashr corrected FDR (lfdr) and the sign of the effect size. The analysis used gene sets from the Human Gene Atlas and PanglaoDB libraries, obtained through GSEApy. Genesets with an FDR q-val less than .1 are shown in figures, all results are in supplemental tables.

#### 5.7.6 Blood cell counts

This category of traits included 31 measures of blood cell counts, percentages, and mean cell volumes (Category 100081).

### 5.8 rDNA variation in GTEx samples

#### 5.8.1 GTEx samples and phenotype data

GTEx v10 sample and subject level metadata were downloaded from AnVIL (dbGaP accession phs000424). Analysis was restricted to subjects for whom rDNA copy-number estimates were successfully obtained after correction (see above). Per-tissue subject counts were determined by collapsing subject IDs and tallying by detailed tissue level (SMTSD).

#### 5.8.2 CNV estimation

5S and 45S copy number variation was determined using the same pipeline described above, starting from GTEx v10 CRAM files mapped to GRCh38.

#### 5.8.3 Differential expression modeling

Differential expression analyses were performed separately for each tissue and for the 45S and 5S rDNA subunits. For each tissue, we modeled transformed gene expression as a function of rDNA copy number and covariates using DESeq2 [49]. To account for technical and unwanted variation, we implemented a principal component-based batch correction strategy. We collected all technical features included in the GTEx sample metadata, plus sex, age, and race. We then encoded categorical variables numerically and performed pairwise absolute Spearman correlations between all covariates using whole blood samples (which are included for nearly every cohort member). Many technical features in the GTEx metadata were highly or perfectly correlated across the cohort (for example, Total Bases and Exonic Reads), so we performed hierarchical clustering on the correlation matrix to select representative metrics from each block of highly correlated attributes. We then performed principal component analysis (PCA) on the matrix of encoded representative features for each tissue and retained the first components that cumulatively explaining more than 90% of the variation in batch. This varied between 3 and 15 PCs, depending on the tissue. These vectors were used to represent covariates in tissue-level DESeq2 designs. Copy number was rank-based inverse normal transformed prior to mod-eling to stablize variance across tissues. For each gene-tissue-subunit combination we obtained log2 fold-changes and and standard errors. For each subunit, we then pooled each set of observations and determined FDR for each observed fold-change using the ashr package in R [50]. Observations were considered significant if they passed 5% FDR.

#### 5.8.4 Pathway enrichment analysis

For each tissue, we used Enrichr in R [51] to find pathways with significant association with copy number. We used the set of detected genes in the GTEx count data as background, and we tested against the following list sources: GO Biological Process 2025, GO Molecular Function 2025, GO Cellular Component 2025, KEGG 2021 Human, Re-actome 2022, and MSigDB Hallmark 2020. We retained results with an adjusted p-value ¡ 0.05. For each significant term, we created a similarity matrix using the Jaccard similarity between gene members of each list. We then built a graph with nodes for each term and edges corresponding to Jaccard similarity, with a threshold of 0.15. We performed Leiden clustering in this graph at a resolution of 0.1 to find a smaller set of lists with high relatedness and selected representative cluster members to plot.

#### 5.8.5 Tissue-level rDNA responsiveness

To quantify how many genes within each tissue were associated with rDNA copy number, we summarized DESeq2 results by tissue. For each tissue we counted genes whose association with 45S or 5S copy number met FDR *<* 2.5% and then classified genes into mutually exclusive categories based on significance and effect direction across subunits:

- “Both, concordant”: 45S and 5S both FDR *<* 0.025 and log(2) fold-changes had the same sign.
- “Both, discordant”: both subunits significant but log(2) fold-changes of opposite sign.
- “45S only”: 45S FDR *<* 0.025 and 5S FDR *>* 0.025.
- “5S only”: 5S FDR *<* 0.025 and 45S FDR *>* 0.025.

#### 5.8.6 Concordance across tissues and subunits

To estimate the concordance in effects between subunits within tissues, we constructed paired log2 fold-change vectors and computed the Spearman correlation across genes. We then extended this analysis across all tissues by concatenating the paired vectors and computing the Spearman correlation. To assess the significance of this concordance for each gene, we randomly permuted the 5S log2 fold-change values across tissues while holding the 45S vector fixed and recomputed the correlation 1000 times to build a null distribution. We considered the bottom quartile of genes, which largely overlapped with the null distribution, to be independent in their subunit effects on gene expression, while the top quartile we considered to be strongly concordant. We then used the R implementation of Enrichr (enrichR) to find biological pathways and GO terms that showed significant enrichment in either group of genes.

#### 5.8.7 Pan-tissue rDNA responsiveness score

To quantify, for each gene, how broadly and consistently expression was associated with rDNA copy number across tissues, we defined a pan-tissue score separately for the 45S and 5S subunits. For a given subunit, we first identified genes significant in at least one tissue at FDR *<* 2.5% and computed, for each such gene:

- the number of tissue with FDR *<* 2.5% = the breadth of the effect
- the fraction of tissues with the same log2 fold-change sign = sign consistency
- the mean log2 fold-change value across tissues
- the maximum −log10(FDR) = the strength of the effect

The pan-tissue score is the product of breadth, sign consistency, and strength

### 5.9 rDNA variation in pancreatic islet microtissues

#### 5.9.1 Islet data

Human pancreatic microtissue organoids and corresponding functional data from 67 distinct donors were procured from InSphero AG (Schlieren, Switzerland). In brief, InSphero obtained primary human islets from Prodo Laboratories (Irvine, USA) from anonymized deceased donors with consent from next of kin. Islets were enzymatically dissociated at 37C using TrypLE Express (Thermo Fisher Scientific, 12604013) supplemented with 40 µg/mL DNase I (Sigma-Aldrich, 10104159001). After removal of undispersed clusters, approximately 2,000 viable single cells were seeded into Akura PLUS 96-well hanging-drop plates to allow scaffold-free reaggregation for 5 days. The resulting microtissues were then transferred to Akura 96-well ultra-low attachment plates and maintained in 3D InSight Human Islet Maintenance Medium. Functional characterization was performed after at least 8 days of maturation from the start of aggregation. Culture medium was removed, and islet microtissues were washed twice with 70 µL 3D InSight Krebs-Ringer-HEPES Buffer (KRHB, InSphero, CS-07-051-01) containing 2.8 mM glucose, followed by a 1-hour equilibration in the same solution. Glucose-stimulated insulin secretion (GSIS) was performed using 50 µL KRHB supplemented with the indicated glucose concentrations for 2 hours. Supernatants were collected, and insulin levels were quantified using the Stellux Chemi Human Insulin ELISA (Alpco, 80-INSHU-CH10) and measured on an Infinite M1000 microplate reader (TECAN, Switzerland). InSphero provided key donor metadata, such as age and sex, which were included as variables in models.

#### 5.9.2 Islet sequencing and associations

WGS libraries were generated from precursor material generated during the microtissue process. We mapped the UMI and non-UMI libraries (60 and 53 samples each) to the GRCh38 genome build, following the same pipeline as used by the 1000 Genomes project [24]. Starting with each UMI / non-UMI aligned bam file, we estimated 5S and 45S CN with the same pipeline used for UK Biobank data (see above).

Using insulin secretion (IS) measurements at baseline glucose levels of 2.8 mM, we tested for the additive and sex-interaction effect of CN on IS, controlling for donor age. Similarly, using IS measurements at all glucose levels, we tested for the effect of CN on IS at each glucose level (treated as a categorical variable). For both tests, we used a linear mixed effects model (`mixedlm` implemented in `statsmodel v0.13.2`), grouping on donor index to allow for dependence of repeat IS measurements for each donor. We conducted the latter test in a sex-stratified manner to get effect sizes in males and females, separately.

#### 5.9.3 Sample Preparation

Frozen pellets were removed from −80C storage and thawed on dry ice. Each pellet was resuspended in 100 µL of 1 X DNA/RNA Shield solution (R1100-250; Zymo Research). Samples were homogenized by pipetting 10x and briefly centrifuged. Samples that were not fully resuspended were gently pulse-vortexed and centrifuged until homogenous. The resulting homogenate for each sample was then aliquoted into three 33 µL portions for subsequent isolation of genomic DNA, RNA, and backup storage.

#### 5.9.4 Genomic DNA Extraction

Genomic DNA (gDNA) was extracted from the 33 µL aliquots using a modified protocol based on the QIAamp Fast DNA Tissue Kit (51104; Qiagen). The lysis was performed using 232 µL of the following lysis mix: 167 µL of Buffer AVE, 40 µL of Buffer VXL, 1 µL of Reagent DX, 20 µL of Qiagen Proteinase K, and 4 µL of RNase A (100 mg/mL). 232 µL of the prepared lysis mix was added to each 33 µL sample. Samples were incubated at 56C for 10 minutes with shaking at 1000 rpm. Following the incubation, 265 µL of Buffer MVL was added to each tube and mixed thoroughly by pipetting or vortexing. The mixture was then applied to the QIAamp spin column. The remaining extraction steps, including the wash steps with AW1 and AW2 buffers, were performed according to the manufacturer’s instructions. All spin-downs were executed at maximum speed 18,000 x g. Genomic DNA was eluted in two sequential steps using 50 µL of Buffer ATE. For the first elution, 50 µL of Buffer ATE was added to the column membrane, incubated for 5 minutes at room temperature, and centrifuged. A second elution using 50 µL of Buffer ATE was performed immediately following the first, with a final centrifugation to maximize DNA yield. Genomic DNA concentration was determined using the Qubit dsDNA High Sensitivity Assay Kit (Q32851; Thermo Fisher Scientific). The integrity and size distribution of the gDNA were assessed using the 5300 Fragment Analyzer (DNF-488; Agilent).

#### 5.9.5 RNA Extraction

Total RNA was extracted from the 33 µL aliquots of the 1 x DNA/RNA shield cell homogenates (prepared as described in Sample Preparation) using the Quick-RNA Miniprep Plus Kit (R1057, Zymo Research). The lysis step was modified: the sample volume was adjusted to 300 µL with 267 µL of 1 X DNA/RNA shield and mixed via pulse vortexing. A 45 µL Proteinase K mixture (15 µL of Proteinase K combined with 30 µL of PK Digestion Buffer) was added to the lysed sample, mixed, and incubated at room temperature for 1.5 hours. Following digestion, 345 µL of RNA lysis buffer was added, and RNA purification proceeded according to the manufacturer’s protocol. RNA was eluted in 55 µL of DNase/RNase-free water and immediately stored at −80C. The quality and concentration of the extracted RNA were assessed using the 5300 Fragment Analyzer (DNF-471; Agilent).

#### 5.9.6 Whole Genome Sequencing

Whole genome sequencing (WGS) libraries were prepared using two distinct methods: a standard non-UMI approach and a UMI-based approach. For the non-UMI libraries, 50 ng of extracted gDNA was used as input for preparation with the Illumina DNA Prep Kit (20060060; Illumina), following the manufacturer’s protocol. These libraries were indexed using the IDT for Illumina DNA/RNA UD Indexes Set A (20025019; Integrated DNA Technologies). For the UMI-based libraries, 50 ng of gDNA was used as input into the NEBNext Ultra II FS DNA Library Prep Kit (E6177L, New England Biolabs), following the manufacturer’s protocol. Adaptors containing Unique Molecular Identifiers (UMIs) were used for ligation. The adaptors used were the NEBNext Multiplex Oligos for Illumina (Unique Dual Index UMI Adaptors DNA Sets 1; E7395S; New England Biolabs). The final non-UMI and UMI-based libraries were quantified, normalized, and pooled. The pooled libraries were sequenced on an Illumina NovaSeq 6000 to generate 150×150 bp paired-end reads.

#### 5.9.7 RNA-seq Library Preparation

No-selection RNA-Seq libraries were constructed using the NEBNext Ultra II Directional RNA Library Prep Kit (E7760L; New England Biolabs) without the polyA selection step. Due to the high concentration of total unselected RNA, two protocol adjustments were made: the input was reduced to 10 ng of total RNA, and the fragmentation time was increased to 20 minutes (from the standard 15 minutes) to ensure efficient fragmentation of ribosomal RNA. During the ligation step, libraries were indexed using Unique Dual Index UMI RNA Adaptors (E7416S; New England Biolabs). The pooled libraries were then sequenced on an Illumina NovaSeq 6000 with 150×150 bp paired-end reads.

## Data Availability

Summary statistics of all associations reported in this publication are available at 10.6084/m9.figshare.30996919. Individual-level 5S and 45S copy numbers will be available as a returned dataset from UK Biobank. Code for estimating rDNA copy number from whole genome sequencing data will be available at github.com/calico/rDNA_CN.

## 6 Acknowledgements

We thank Burcak Yesildag (InSphero) for expertise on primary human islet biology and assistance with methods documentation. We are grateful to James Lee for guidance on type 2 diabetes and pancreas biology, and to Nick Bernstein for foundational discussions that shaped this work. This research was conducted using the UK Biobank Resource (Application 18448 and 44584). The Genotype-Tissue Expression (GTEx) Project was supported by the Common Fund of the Office of the Director of the National Institutes of Health, and by NCI, NHGRI, NHLBI, NIDA, NIMH, and NINDS.

## 7 Author Contributions

Conceptualization, formal analysis, and writing of the original draft were performed by D.G.H., M.H., A.R., J.S.B., and N.H.T. Methodology was developed by D.G.H., M.H., A.R., J.S.B., N.H.T., N.F., and I.L. MRI-derived phenotypes were generated and quality checked by E.P.S and M.T. All authors contributed to review and editing of the manuscript.

## 9 Data and software availability

Summary statistics of all associations reported in this publication are available at 10. 6084/m9.figshare.30996919. Individual-level 5S and 45S copy numbers will be available as a returned dataset from UK Biobank. Code for estimating rDNA copy number from whole genome sequencing data will be available at github.com/calico/rDNA_CN.

## 10 Supplementary Figures

**Figure 1:**
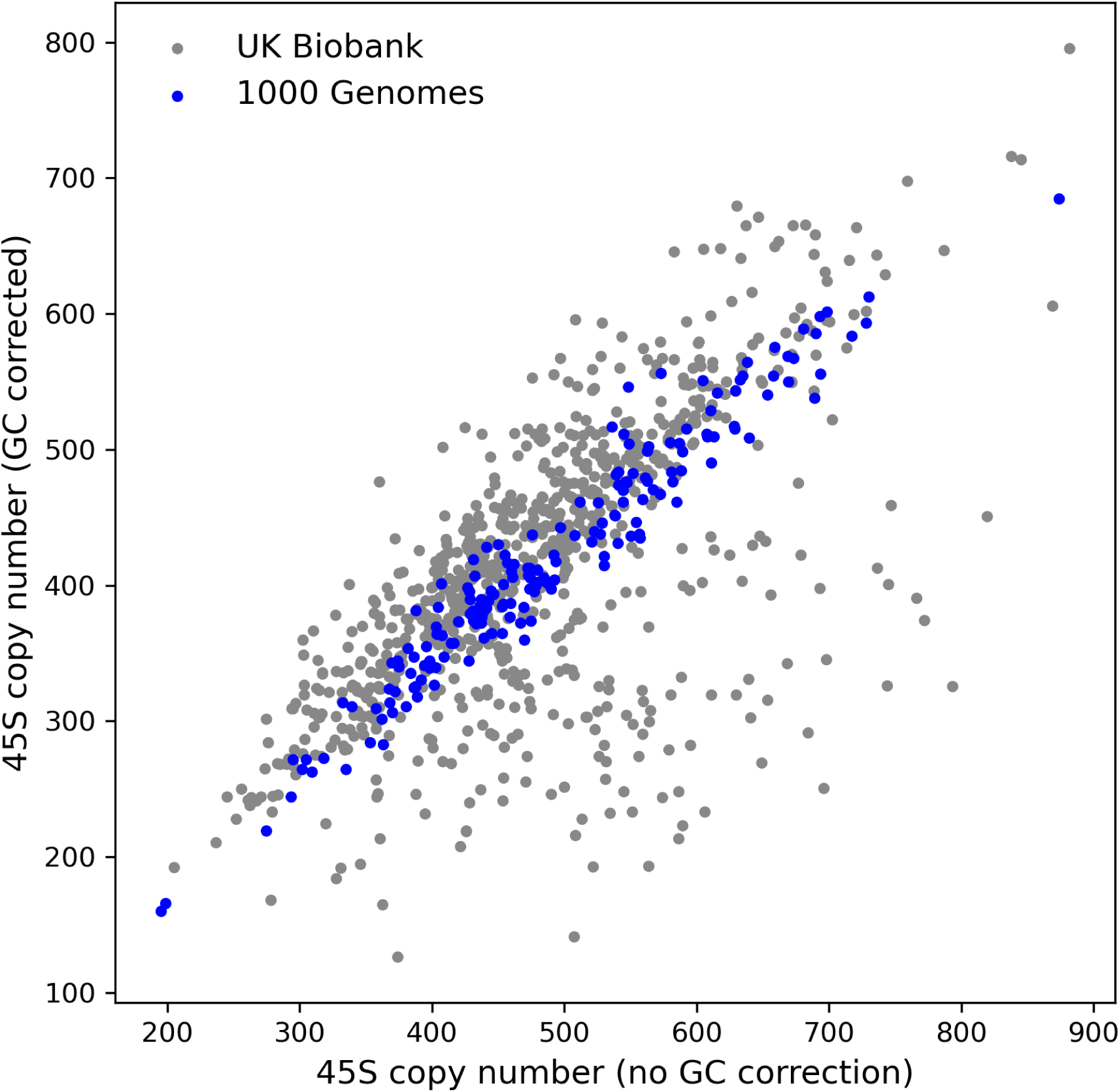
Impact of sequence GC-content on rDNA copy number estimates in two human cohorts.

**Figure 2:**
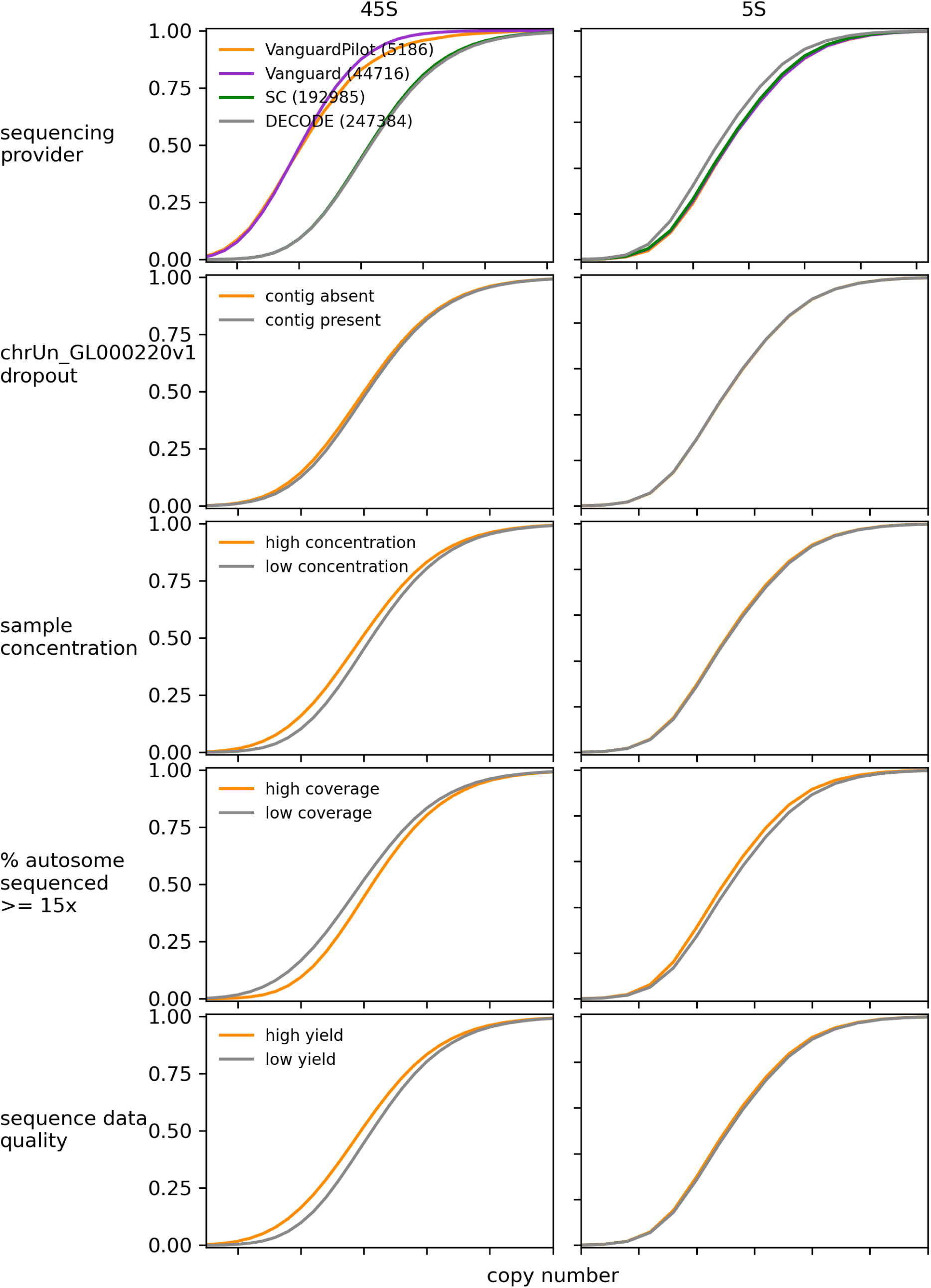
Impact of WGS batch effects on GC-corrected rDNA copy number.

**Figure 3:**
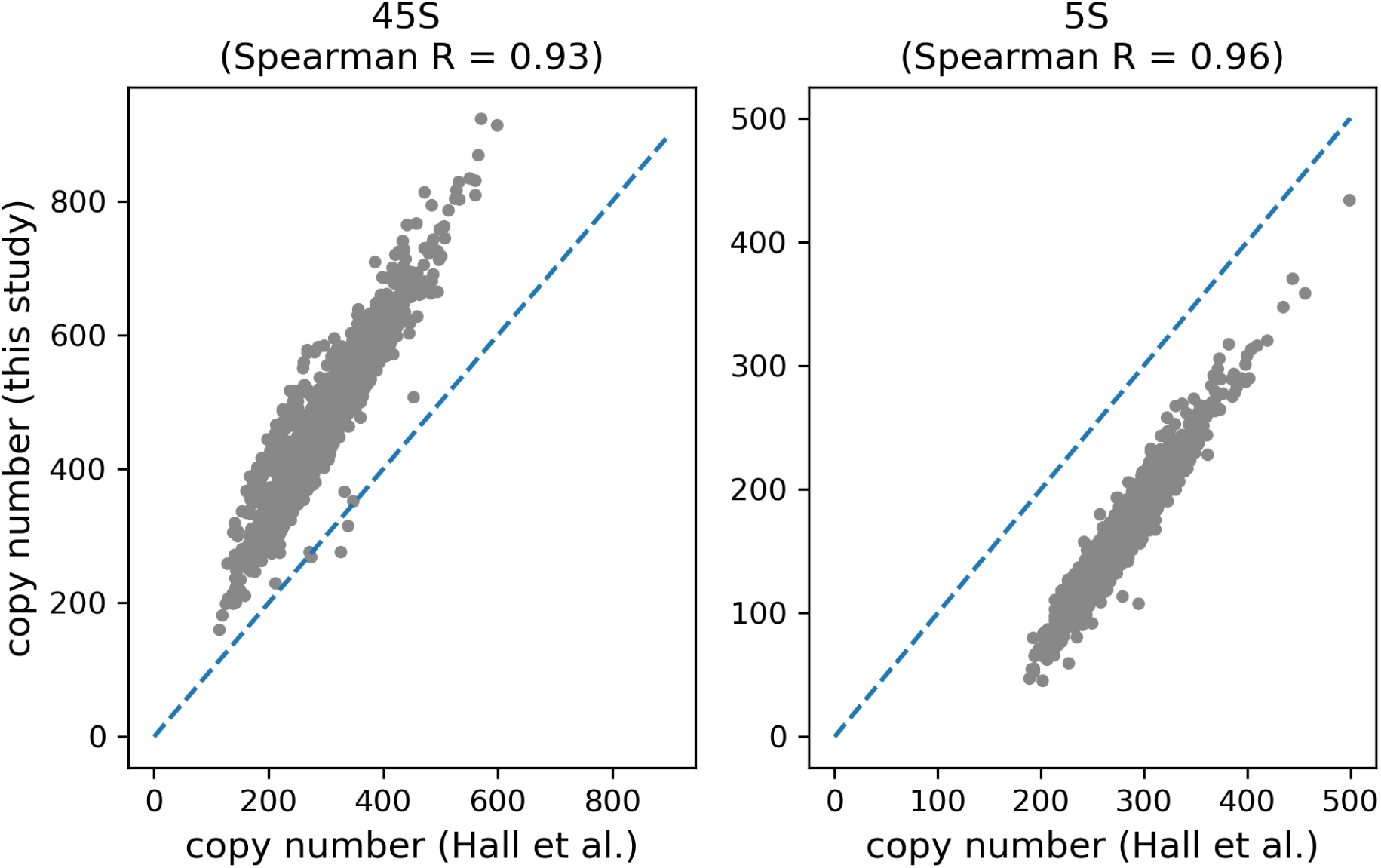
Comparison of 5S and 45S copy number estimates in the 1000 Genomes cohort, between this study and those previously reported by Hall et al.

**Figure 4:**
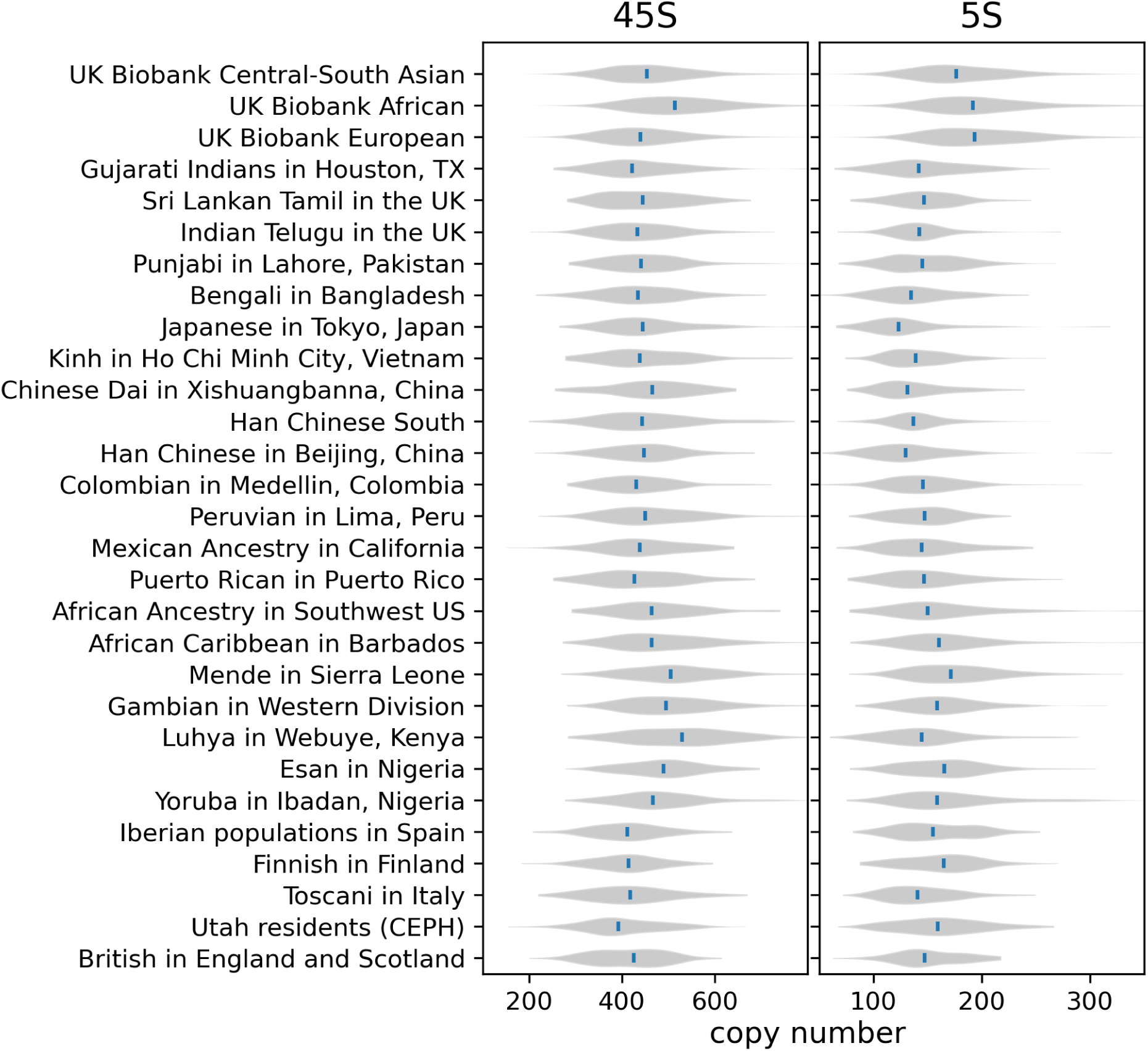
Population distribution of 45S and 5S copy number across 1000 genomes ethnicity.

**Figure 5:**
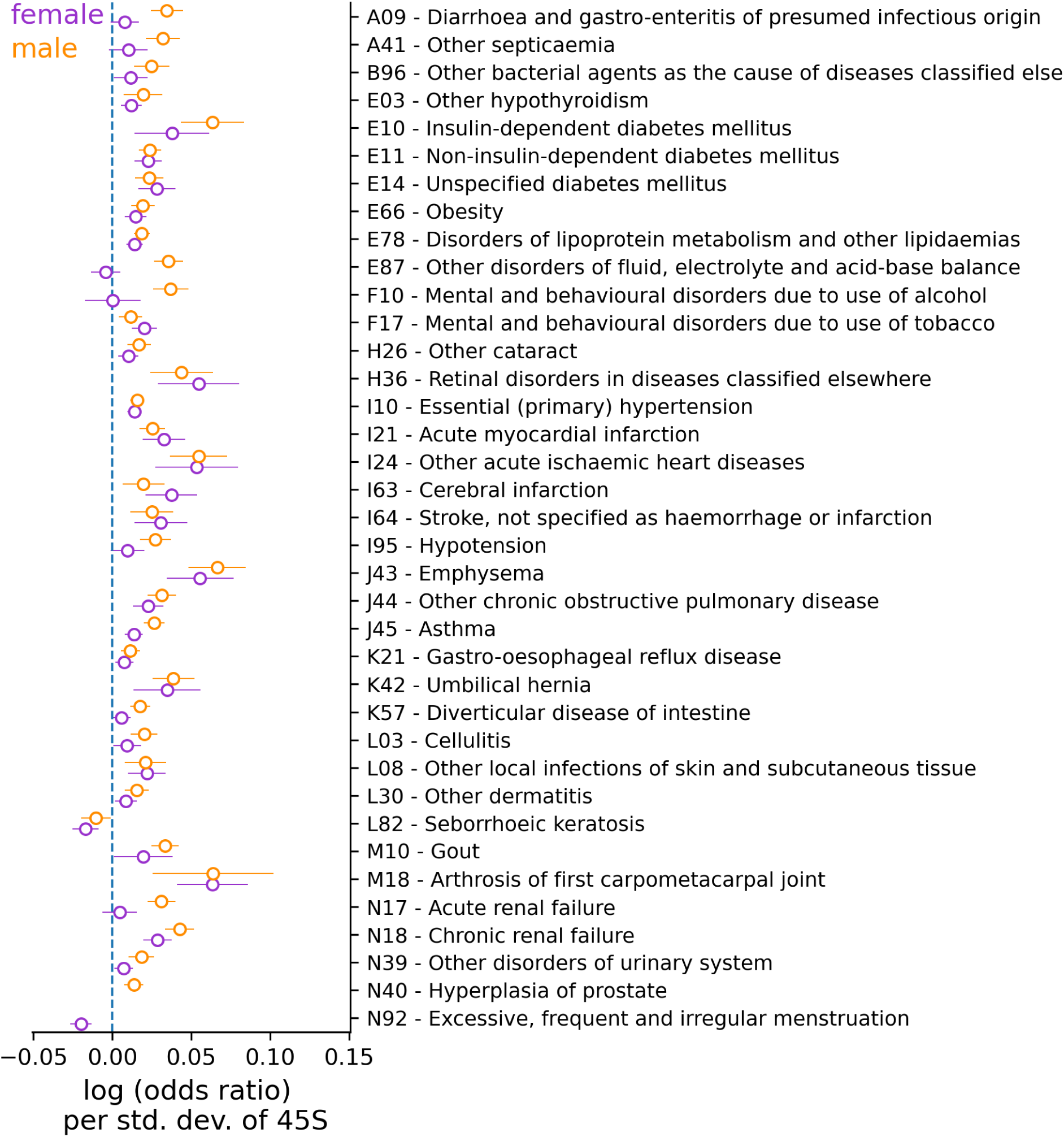
Sex-dependent effect of 45S copy number on disease incidence.

**Figure 6:**
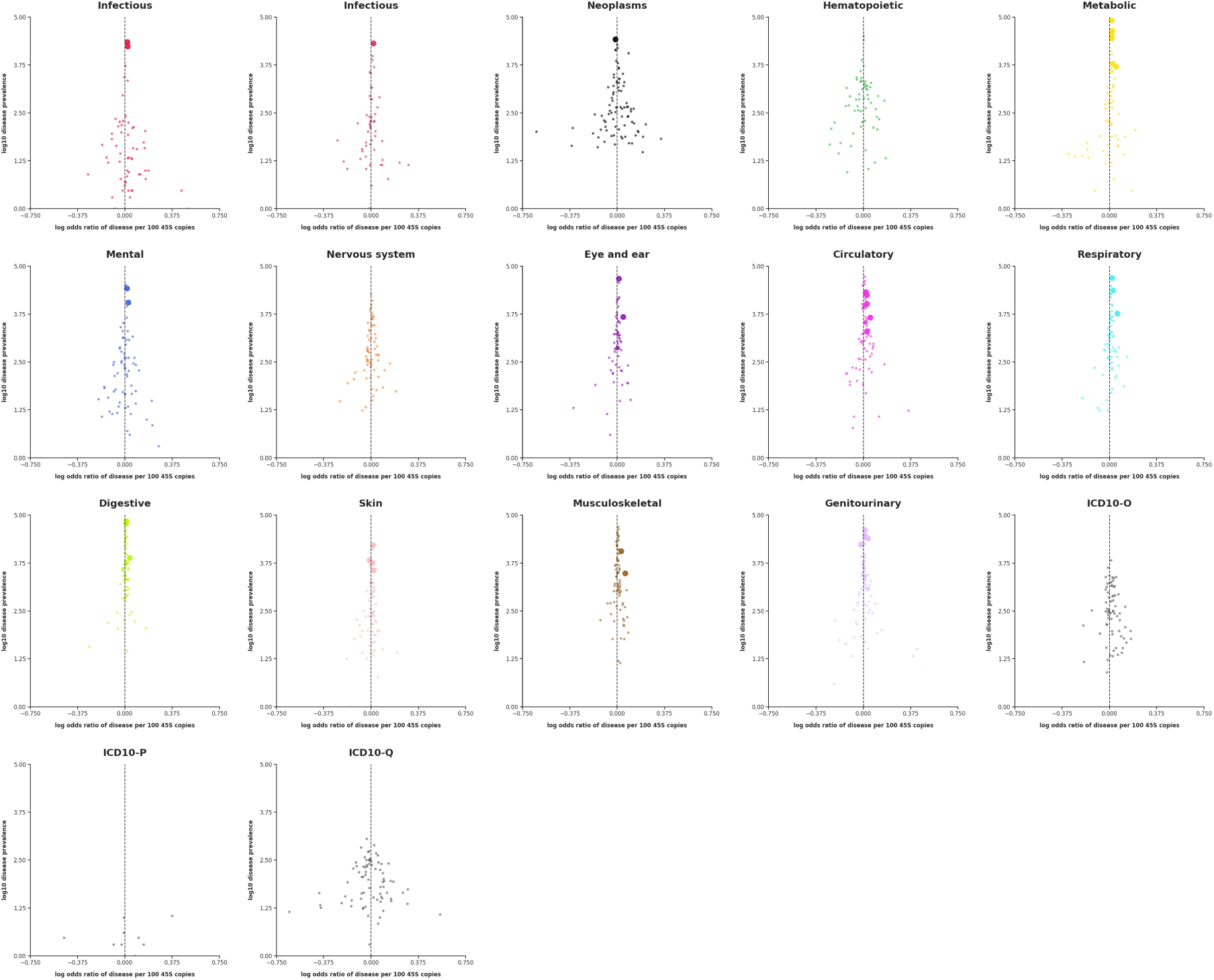
UKBB disease classification incidence.

**Figure 7:**
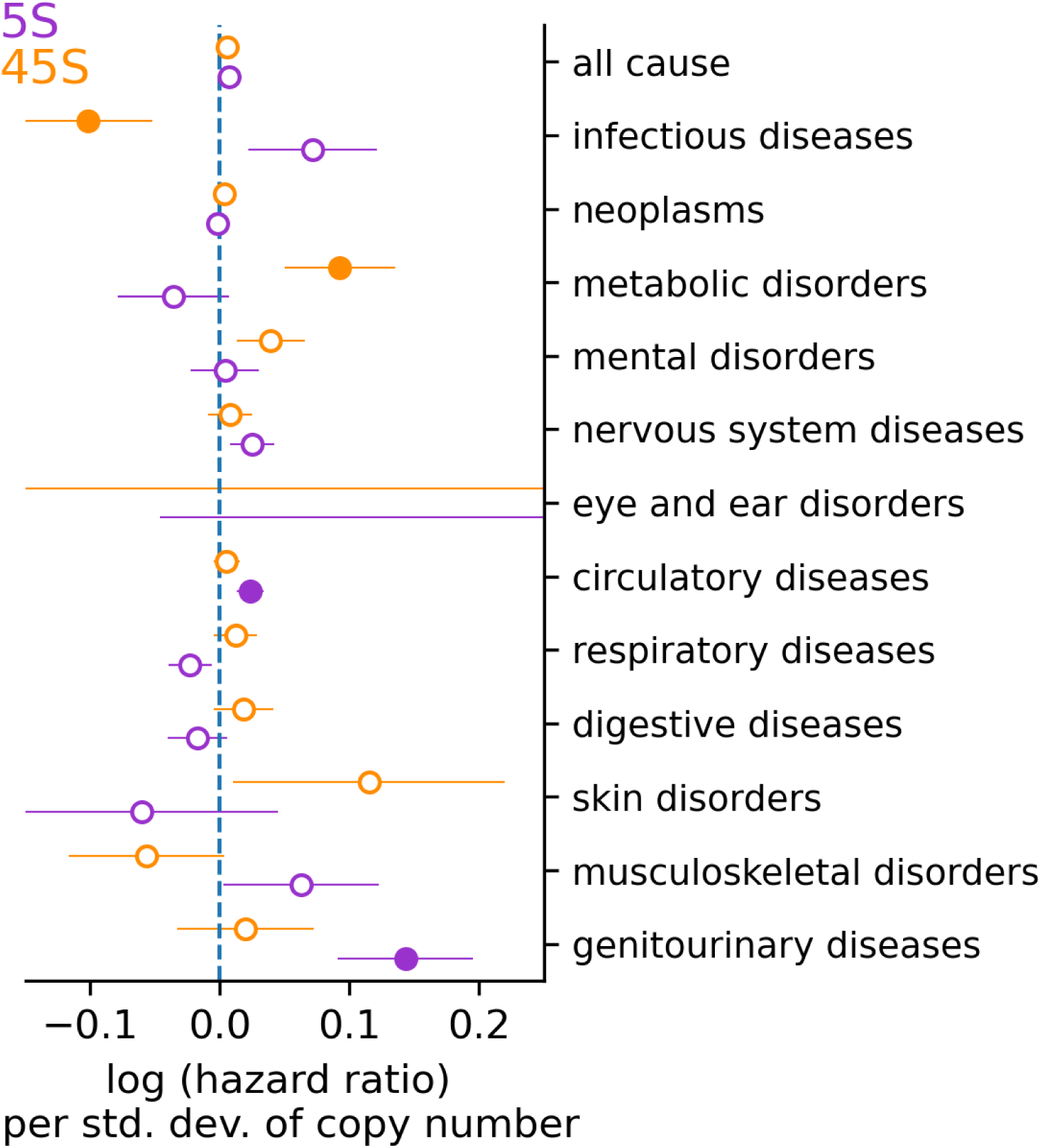
Effect of 5S and 45S copy number on mortality.

**Figure 8:**
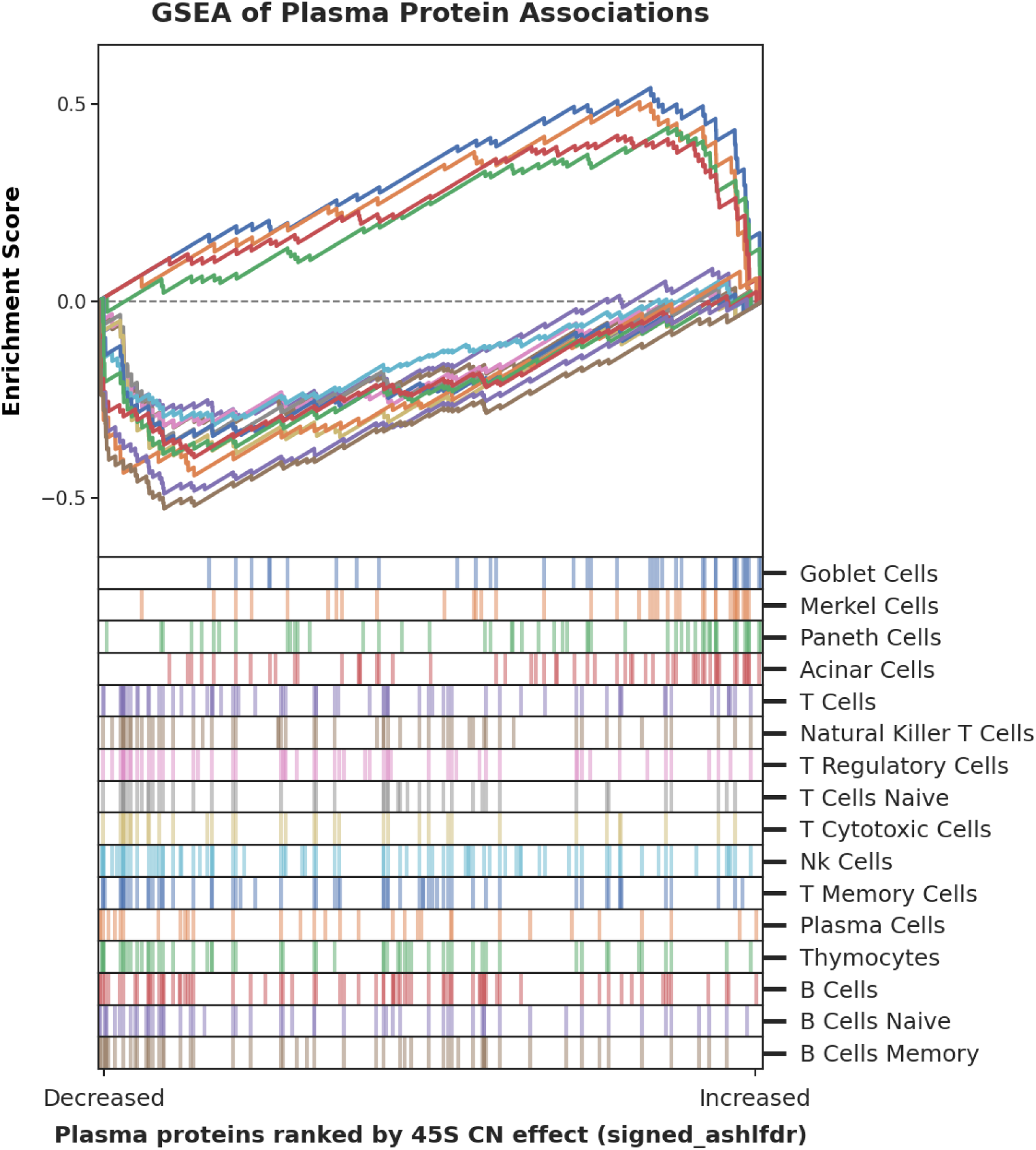
PPP GSEA with PangloDB Gene set.

**Figure 9:**
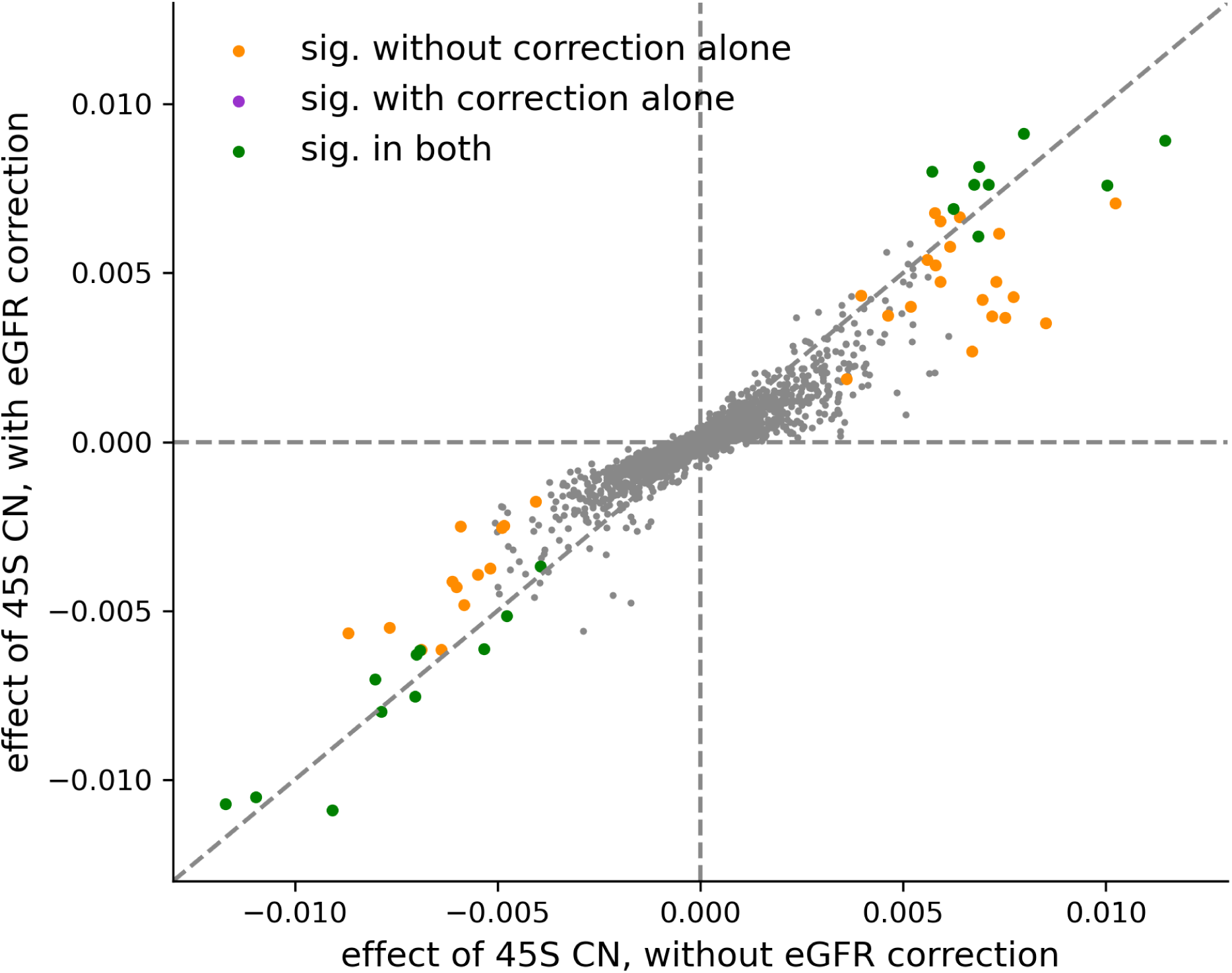
Comparison of the effect of 45S copy number on the plasma proteome, with and without correcting for eGFR levels.

**Figure 10:**
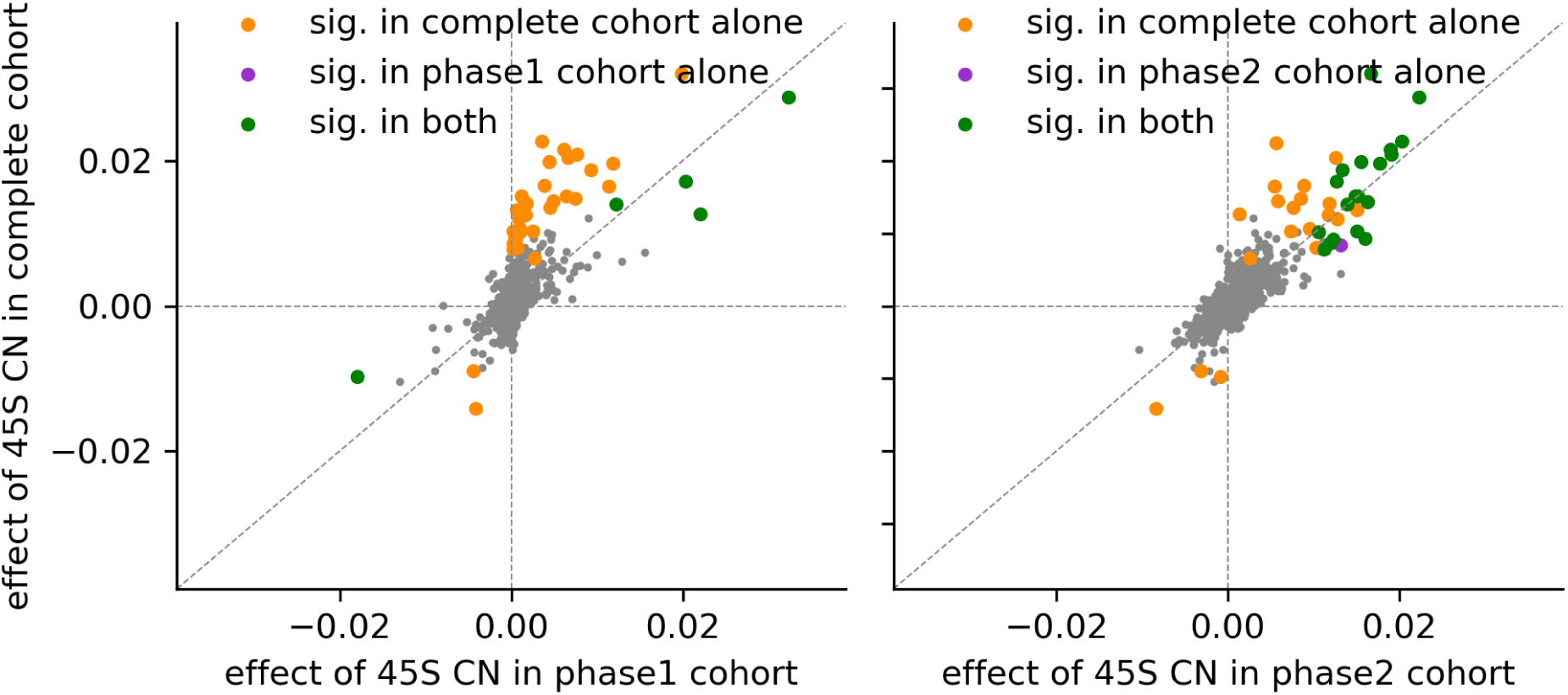
Comparison of the effect of 45S copy number on disease incidence, using the complete cohort and phase1 and phase2 cohorts each.

**Figure 11:**
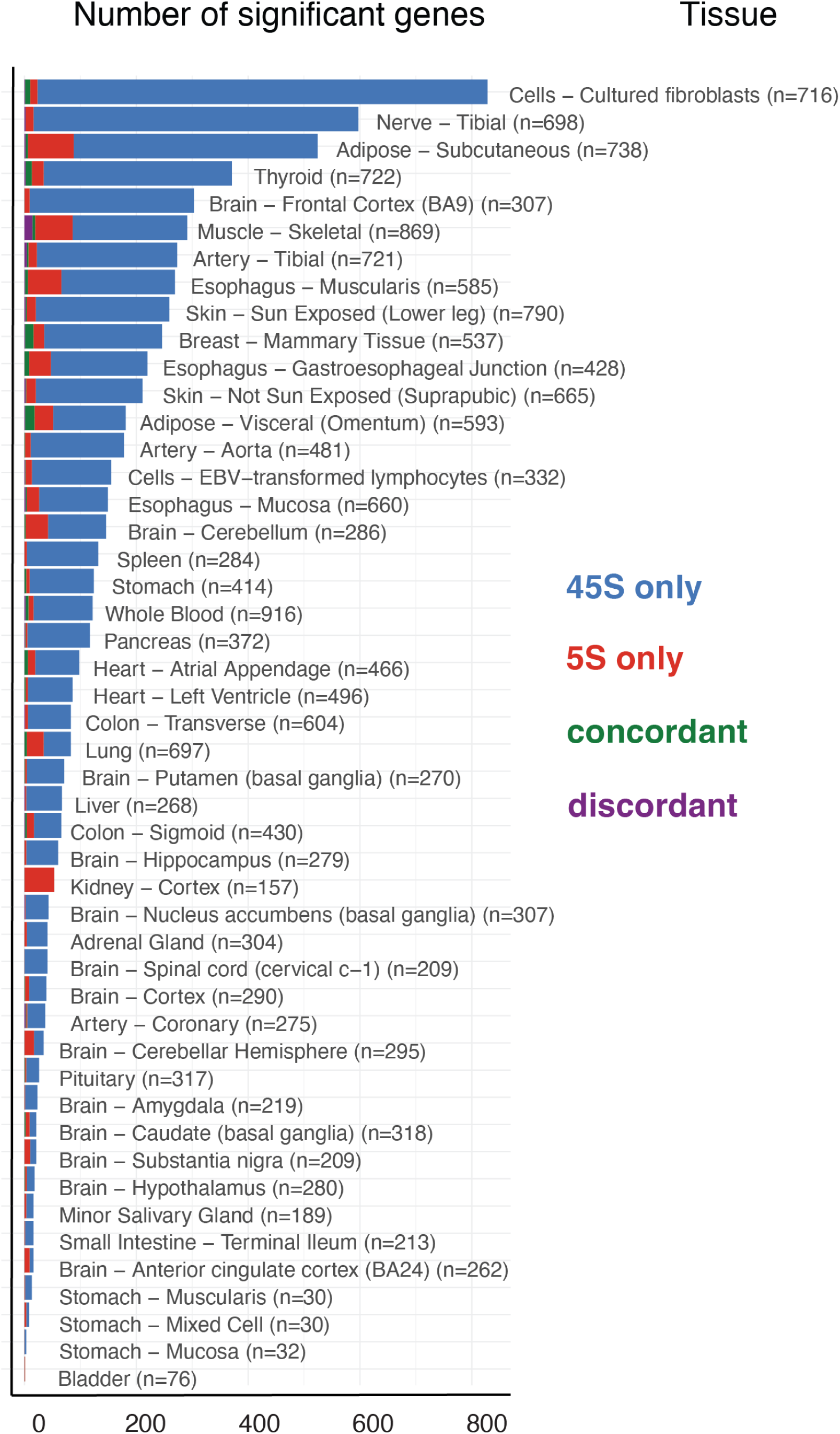
Significant genes per GTEx tissue and array.

**Figure 12:**
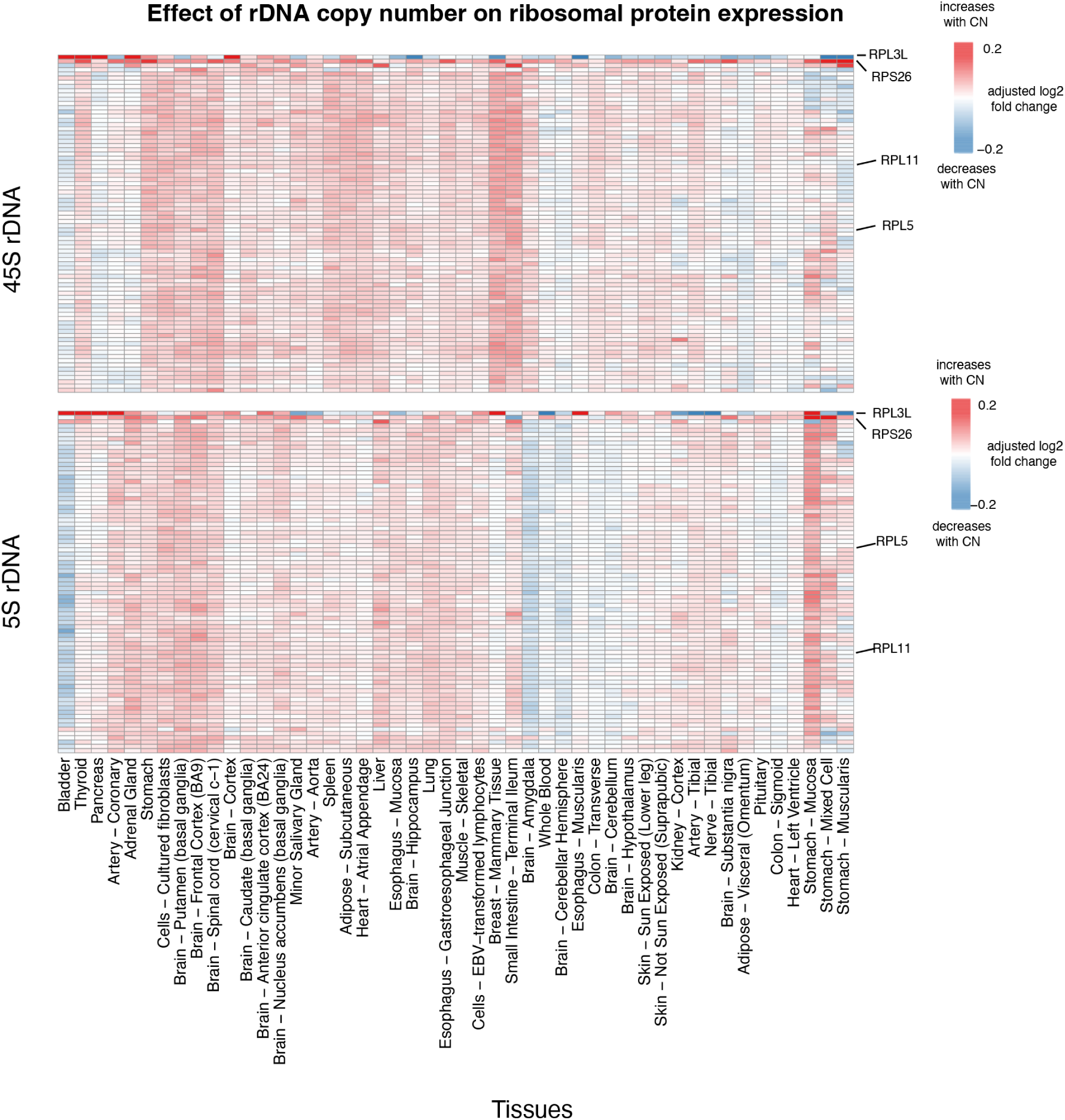
Effect of rDNA copy number on ribosomal protein expression.

**Figure 13:**
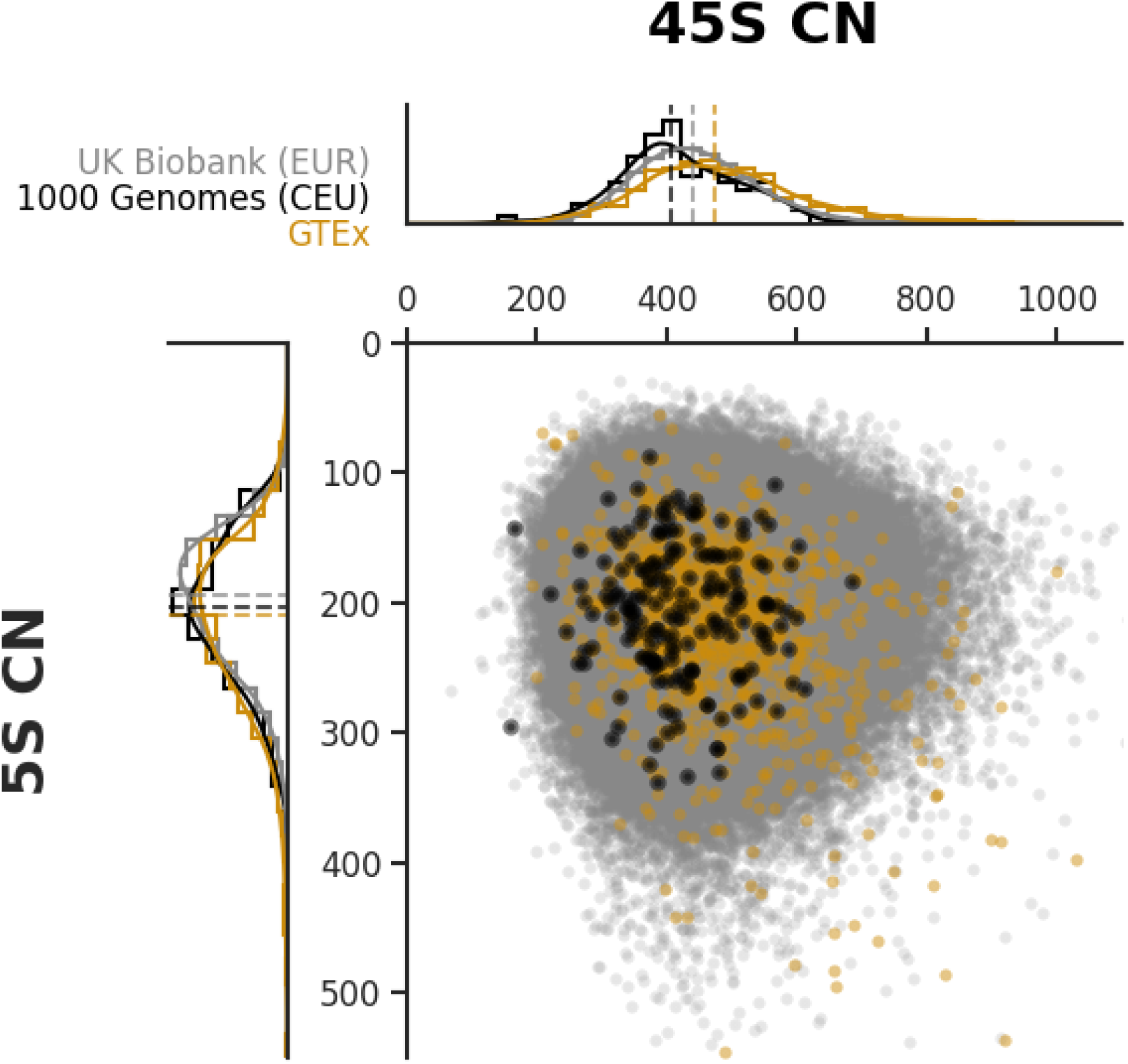
45S rDNA vs 5s rDNA across cohotrs.

